# Missed opportunities for HIV testing among those who accessed sexually transmitted infection (STI) services, tested for STIs and diagnosed with STIs: a systematic review and meta-analysis

**DOI:** 10.1101/2022.06.24.22276855

**Authors:** Kanwal Saleem, Ee Lynn Ting, Andre J.W. Loh, Rachel Baggaley, Maeve B. Mello, Muhammad S. Jamil, Magdalena Barr-Dichiara, Cheryl Johnson, Sami L. Gottlieb, Christopher K. Fairley, Eric P.F. Chow, Jason J. Ong

## Abstract

**Introduction:** Of 37.7 million people living with HIV in 2020, 6.1 million still do not know their HIV status. We synthesise evidence on concurrent HIV testing among people who tested for other sexually transmitted infections (STI).

**Methods:** We conducted a systematic review using five databases, HIV conferences and clinical trial registries. We included publications between 2010 and May 2021 that reported primary data on concurrent HIV/STI testing. We conducted a random-effects meta-analysis and meta-regression of the pooled proportion for concurrent HIV/STI testing.

**Results and discussion:** We identified 96 eligible studies: the majority (73%) were from high-income countries (HIC), with a third from general populations (36%) and non-heterosexual populations (30%). Among the 96 studies, 18 studies had relevant data for a meta-analysis for the proportion of people tested for HIV among those attending a clinic with STI testing services, 15 studies among those tested for other STIs, 13 studies among those diagnosed with STI and three studies for people with STI symptoms. The remaining studies provided data on the acceptability, feasibility, barriers, facilitators, economic evaluation and social harms of concurrent HIV/STI testing. The pooled proportion of people tested for HIV among those attending an STI service (n=18 studies) was 71.0% [95% confidence intervals: 61.0-80.1, I2=99.9%], people tested for HIV among those who were tested for STIs (n=15) was 61.3% [53.9-68.4, I2=99.9%], people tested for HIV among those who were diagnosed with an STI (n=13) was 35.3% [27.1-43.9, I2=99.9%]. and people tested for HIV among those presenting with STI symptoms (n=3) was 27.1% [20.5-34.3, I2=92.0%]. The meta-regression analysis found that heterogeneity was driven mainly by identity as a sexual minority, the latest year of study, country-income level and region of the world.

**Conclusions:** Not testing for HIV amongst people using STI services presents a significant missed opportunity, particularly among those diagnosed with an STI. Stronger integration of HIV and STI services is urgently needed to improve prevention, early diagnosis, and linkage to care services.

**PROSPERO Number:** CRD42021231321

## Introduction

According to the Joint United Nations Programme on HIV/AIDS (UNAIDS), approximately 37.7 million people were living with HIV (PLHIV) in 2020, including 36 million adults and 1.7 million children; of these, 6.1 million people globally were not aware of their HIV status [1]. Access to early testing is essential for HIV prevention, treatment, and linkage to care. Earlier diagnosis and subsequent antiretroviral therapy (ART) initiation significantly decrease HIV-related morbidity and mortality and the risk of onward transmission, resulting in the improved long-term health of PLHIV and their communities [2]. Therefore, more efficient and effective ways to reach the UN global targets to diagnose 95% of PLHIV and link them to care are required. To achieve this, countries need to develop a strategic mix of testing approaches; this can include targeted testing based on risk and symptoms [3] and routine testing for people attending clinical services for sexually transmitted infections (STIs).

Individuals with STIs are at an increased risk of transmitting and acquiring HIV due to biological factors and similar high-risk sexual practices such as condomless sex or multiple sexual partners [4]. Studies show that detecting and treating STIs may reduce HIV acquisition and transmission [5]. These findings underscore the need for improved routine STI services that include the offer of HIV testing among people tested for STIs (i.e., concurrent HIV/STI testing) at the same visit. Regular concurrent HIV/STI testing for those at higher risk facilitates early HIV diagnosis and might also reduce the onward transmission of HIV or other STIs [6].

In 2007, the World Health Organization (WHO) recommended the routine offer of concurrent HIV testing in all STI services[7] and reinforced this in all subsequent testing guidelines. In 2019, there were further recommendations for testing and retesting for people presenting with a diagnosis or receiving treatment for STIs [8]. This includes dual HIV/syphilis rapid diagnostic tests that can be considered the first test in HIV testing strategies and algorithms in antenatal care settings. HIV testing is also recommended to be integrated with other clinical services, including STIs and tuberculosis, to create opportunities for the early diagnosis of co-infections and increase uptake of HIV testing among populations at higher risk for HIV infection [9].

Despite these long-standing global guidelines, HIV testing among people tested for STIs or presenting with STI symptoms in diverse healthcare settings (community-based services, hospitals, STI clinics, and physician/primary care outpatient clinics) remains suboptimal. In 2016, a retrospective USA-based study with participants from 29 states showed that only 43% of the participants diagnosed with an STI in a physician outpatient clinic or emergency department were screened for HIV [10]. Similarly, a Spain-based study conducted in 2016 reported that HIV testing was conducted among 61% of people diagnosed with other STIs in various settings, including primary care, hospital or clinic, sexual health clinic and medical specialist [11]. Furthermore, data from the paediatric department of Cincinnati Children’s Hospital Medical Centre in the USA observed test uptake as low as 3.6% among adolescents diagnosed with an STI [12].

A prior systematic review identified HIV testing interventions among health care settings in Europe[13] and another on how incentives could improve HIV/STI testing rates [14]. However, to our knowledge, there are no systematic reviews on concurrent HIV/STI testing uptake across different healthcare settings globally. This systematic review and meta-analysis aim to synthesise the existing evidence on the routine offer and uptake of HIV testing among people attending an STI service, tested for other STIs, diagnosed with STIs or with STI symptoms. Secondary aims included identifying barriers and facilitators for concurrent HIV/STI testing.

## Methods

We conducted a systematic review (Prospero: CRD42021231321) that followed the guidelines in the Cochrane handbook for systematic reviews[15] and the PRISMA (Preferred reporting items for systematic reviews and meta-analyses) guidelines for reporting [16]. Five databases (Ovid MEDLINE, Ovid Embase, Ovid Global Health, EBSCO CINAHL Plus and Web of Science Core Collection), conferences related to HIV (Conference on Retroviruses and Opportunistic Infections (CROI), HIV Research for Prevention conference (HIVR4P), International AIDS Conference (IAS), International Conference on AIDS and STIs in Africa (ICASA), US CDC Prevention, British Association for Sexual Health and HIV (BASHH) and clinical trial registries (clinicaltrials.gov, WHO international clinical trials registry platform) were searched for publications between 1^st^ January 2010 and 5^th^ May 2021 that documented primary data on concurrent HIV/STI testing uptake.

### Study eligibility criteria

We included any studies in English and contained data for the number of people tested for HIV among the number of people who attended an STI service, tested for STIs, diagnosed with STIs (chlamydia, gonorrhoea or syphilis) or with symptoms of STIs. As part of our secondary outcomes, we also included studies reporting the acceptability, feasibility, barriers, facilitators, economic evaluation and social harms of concurrent HIV-STI testing. We excluded duplicated results from the same study or laboratory studies testing HIV diagnostic performance.

### Search method and data extraction

Key concepts included in the search strategy were: (1) HIV and STIs; (2) tests and screening; and (3) early diagnosis, missed opportunities. Additional details on the search strategy are included in the Appendix. All studies’ titles and abstracts were independently screened by at least two reviewers (KS, ET, AL) using the inclusion criteria. The full texts of potentially relevant papers were independently screened by at least two reviewers (KS, ET, AL) and any discrepancies were resolved by another researcher (JO). Relevant data related to primary and secondary outcomes were extracted from deduplicated publications. We conducted a qualitative synthesis of factors associated with concurrent HIV/STI testing and classified each attribute using the socio-ecological model[17]: “Individual factors”, “service factors”, and “societal factors”.

### Statistical analysis

Random-effects meta-analysis was used to calculate across-study pooled proportions of people tested for HIV among those attending an STI service, tested for other STIs or diagnosed with STIs. Pooled proportions and 95% confidence intervals were generated using a Freeman-Tukey–type double arcsine transformation to adjust for variance instability. Statistical heterogeneity between studies was assessed with the *I*^2^ statistic. Predefined subgroup meta-analyses were based on the following covariates: country-income level, type of HIV testing (rapid testing, venepuncture), recruitment site, study population, the latest year of study and region of the world. Funnel plots were generated to assess the possibility of small-study effects associated with publication bias. Egger’s test was performed to confirm the presence of this bias. When publication bias was significant (p<0.05), we used a nonparametric trim-and-fill analysis to explore the sensitivity of the meta-analysis results to potentially omitted studies. Random-effects meta-regression models using the covariates described above were conducted to examine the association of these variables with the effect size. Adjusted R^2^ is reported for the percentage of variance explained by the subgroups above. All analyses were conducted using Stata, version 17.0 (StataCorp LLC). We evaluated the methodological quality using the Cochrane risk of bias tool for randomised controlled trials, Newcastle-Ottawa quality assessment scale for cross-sectional, cohort and case-control studies, consolidated health economic evaluation reporting standards (CHEERS) checklist for economic evaluation studies, and Joanna Briggs Institute (JBI) critical appraisal checklist for qualitative studies.

## Results and discussion

Of 7582 articles, 612 full texts were examined, and 96 studies were included in our final analysis (Figure 1). Among the 96 studies, 18 studies had relevant data for a meta-analysis for the proportion of people tested for HIV among those attending an STI service, 15 studies among those tested for STIs, 13 studies among those diagnosed with STI, and three studies for people with STI symptoms. The remaining studies provided data related to the secondary outcomes. Table 1 provides an overview of the included studies. In brief, the majority (73%) of studies were from HIC, with about a third from general populations (36%) and sexual minorities (i.e., non-heterosexual population) (30%). (Supplementary Table 1 provides further details of each included study.)

**Table 1.**
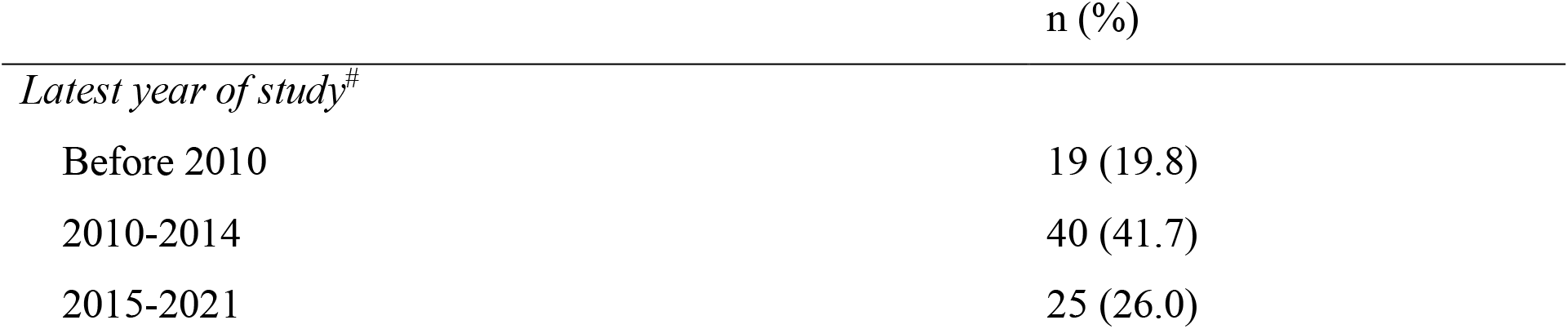
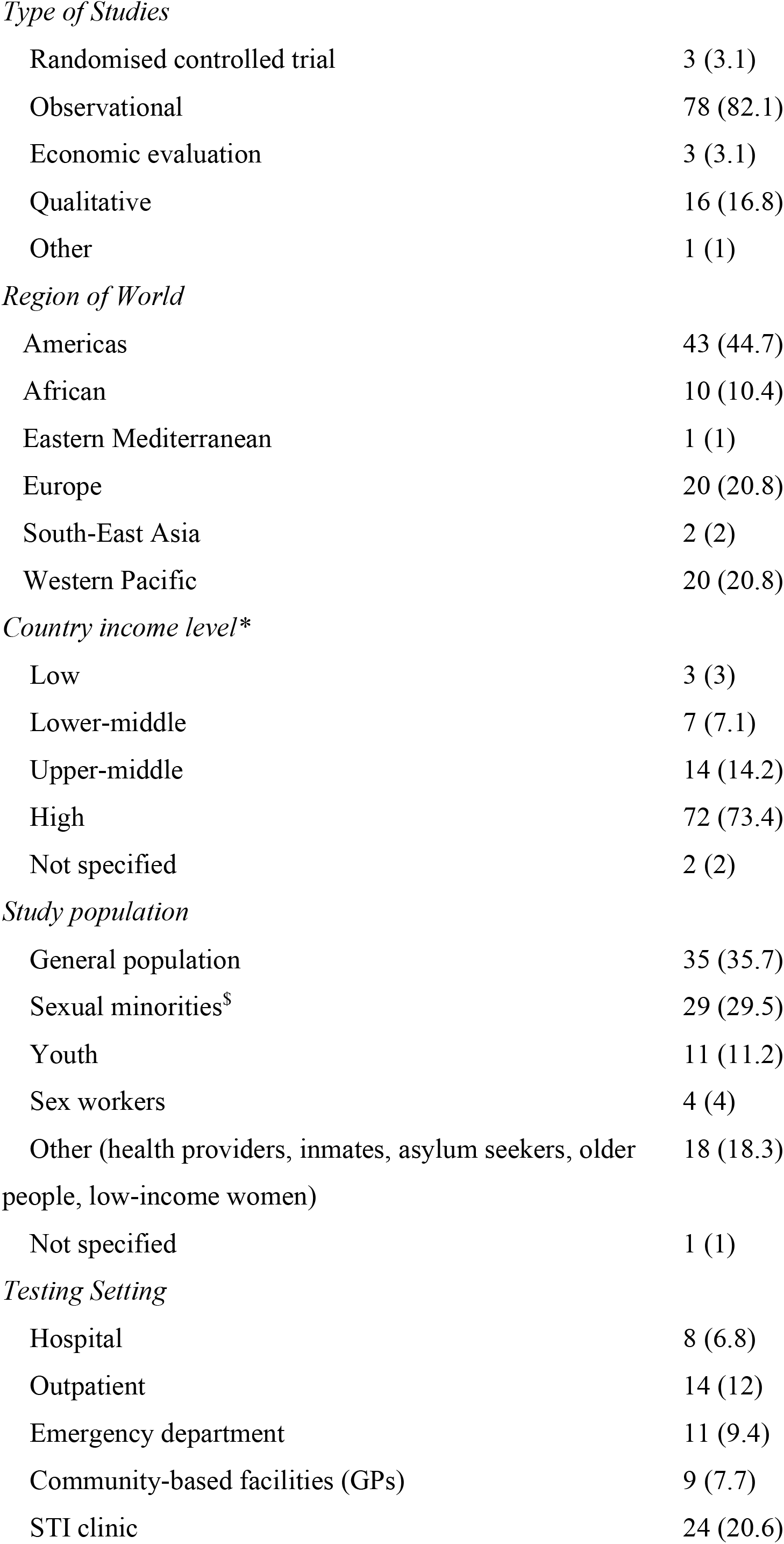
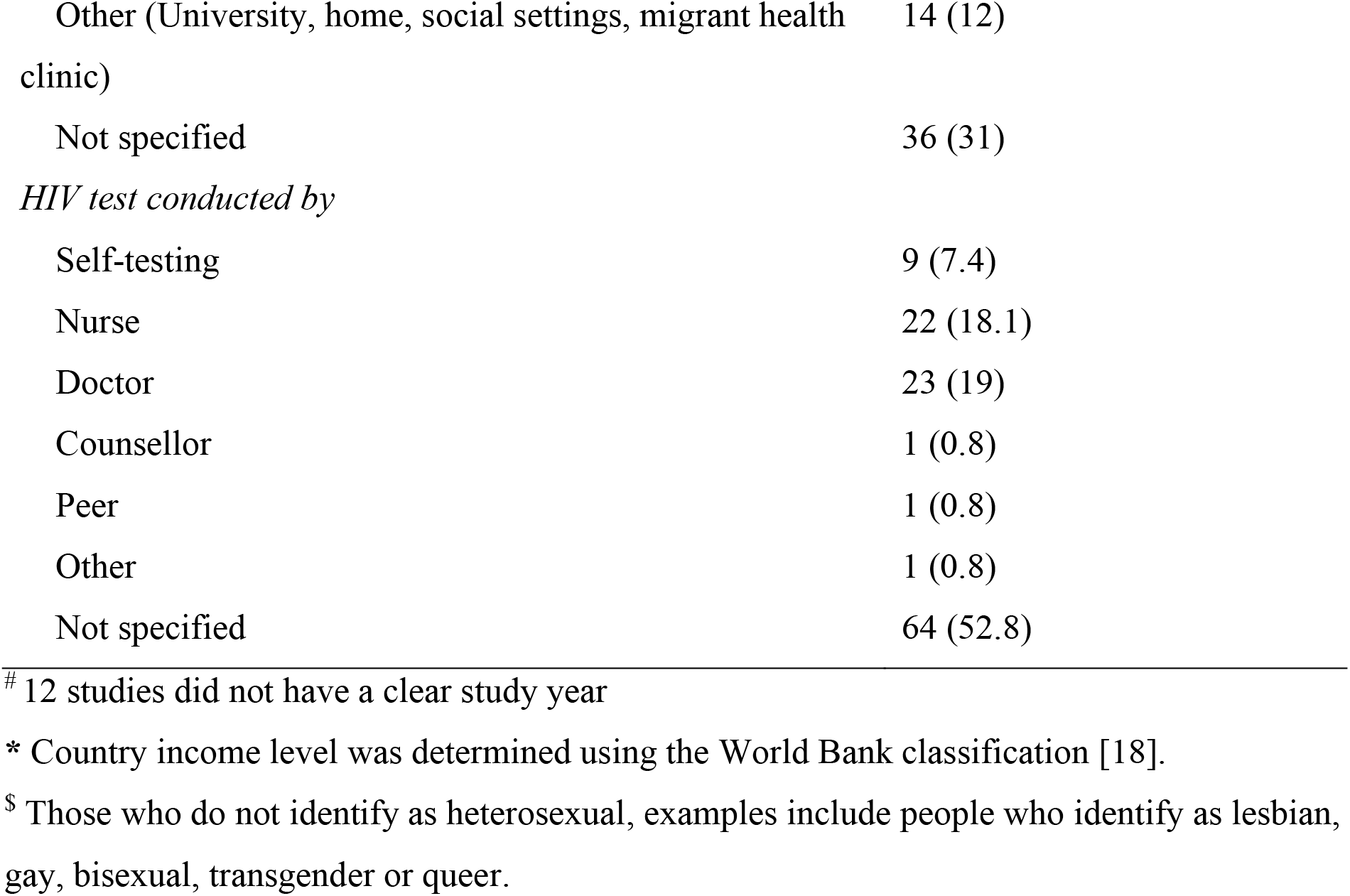
Summary of included studies (N=96)

|  | n (%) |
| --- | --- |
| *Latest year of study*<sup>#</sup> |  |
| Before 2010 | 19 (19.8) |
| 2010-2014 | 40 (41.7) |
| 2015-2021 | 25 (26.0) |
| *Type of Studies* |  |
| Randomised controlled trial | 3 (3.1) |
| Observational | 78 (82.1) |
| Economic evaluation | 3 (3.1) |
| Qualitative | 16 (16.8) |
| Other | 1 (1) |
| *Region of World* |  |
| Americas | 43 (44.7) |
| African | 10 (10.4) |
| Eastern Mediterranean | 1 (1) |
| Europe | 20 (20.8) |
| South-East Asia | 2 (2) |
| Western Pacific | 20 (20.8) |
| *Country income level\** |  |
| Low | 3 (3) |
| Lower-middle | 7 (7.1) |
| Upper-middle | 14 (14.2) |
| High | 72 (73.4) |
| Not specified | 2 (2) |
| *Study population* |  |
| General population | 35 (35.7) |
| Sexual minorities<sup>\$</sup> | 29 (29.5) |
| Youth | 11 (11.2) |
| Sex workers | 4 (4) |
| Other (health providers, inmates, asylum seekers, older people, low-income women) | 18 (18.3) |
| Not specified | 1 (1) |
| *Testing Setting* |  |
| Hospital | 8 (6.8) |
| Outpatient | 14 (12) |
| Emergency department | 11 (9.4) |
| Community-based facilities (GPs) | 9 (7.7) |
| STI clinic | 24 (20.6) |
| Other (University, home, social settings, migrant health clinic) | 14 (12) |
| Not specified | 36 (31) |
| *HIV test conducted by* |  |
| Self-testing | 9 (7.4) |
| Nurse | 22 (18.1) |
| Doctor | 23 (19) |
| Counsellor | 1 (0.8) |
| Peer | 1 (0.8) |
| Other | 1 (0.8) |
| Not specified | 64 (52.8) |

**Figure 1.**
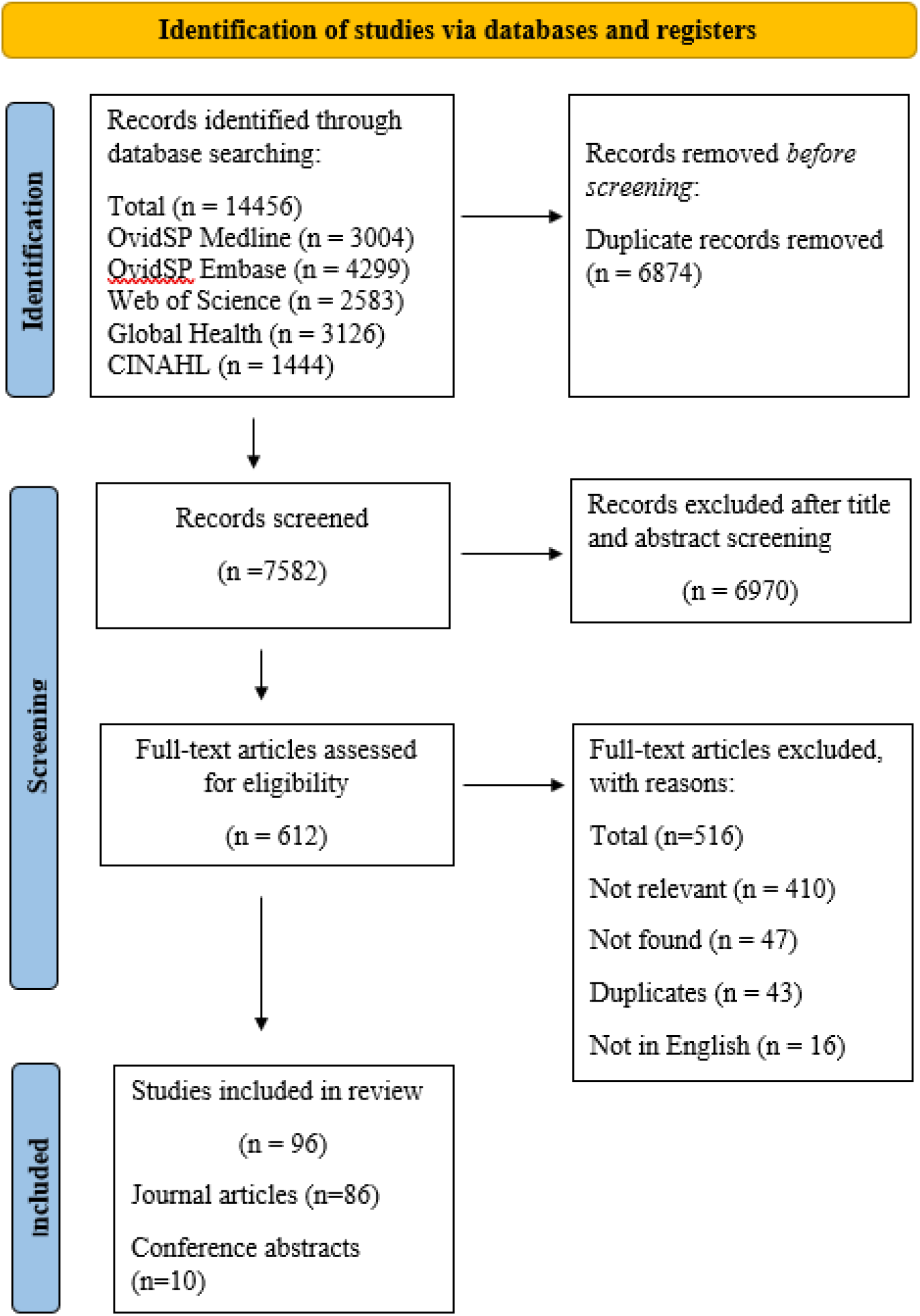
PRISMA flowchart.

### Percentage of people tested for HIV among those attending a clinic with STI testing service

Eighteen studies provided 21 estimates for the meta-analysis (Figure 2, Table 2). The pooled percentage of people tested for HIV when attending an STI service was 71.0% (95% CI: 61.0-80.1%). There was no evidence of publication bias (p=0.837) (Supplementary Figure 1). The meta-regression analysis revealed that sexual minority populations compared with those not belonging to a sexual minority (AOR 1.37, 95%CI:1.10-1.71) and recent latest year of study (2010-2014: AOR 1.68 (95%CI: 1.17-2.40); 2015 onwards: AOR 1.46 (95%CI:1.26-1.29) compared with before 2010, explained most of the heterogeneity (adjusted R^2^=91.3%) (Supplementary Table 2)

**Table 2.** Pooled percentage of people tested for HIV who attended an STI service.

| | Number<br>of studies | Pooled % of people tested<br>for HIV | $I^2$ (p value) |
| --- | --- | --- | --- |
| Total | 18 | 71.0 (61.0-80.1) | 99.95% (<0.001) |
| Country income level |  |  |  |
| High | 11 | 75.4 (66.3-83.4) | 99.93% (<0.001) |
| Middle | 7 | 63.5 (46.4-79.0) | 99.89% (<0.001) |
| Type of HIV testing |  |  |  |
| Rapid HIV testing | 4 | 74.5 (52.6-91.2) | 99.85% (<0.001) |
| Venepuncture | 4 | 62.1 (61.0-80.1) | 99.88% (<0.001) |
| Unclear | 10 | 74.3 (60.5-86.0) | 99.97% (<0.001) |
| Study population |  |  |  |
| Sexual minorities | 7 | 79.1 (70.8-86.3) | 99.73% (<0.001) |
| Not sexual minorities | 11 | 64.5 (46.5-80.5) | 99.97% (<0.001) |
| Latest year of study |  |  |  |
| Before 2010 | 6 | 52.3 (40.3-64.1) | 99.79% (<0.001) |
| 2010-2014 | 6 | 76.9 (64.4-87.3) | 99.95% (<0.001) |
| 2015 onwards | 6 | 88.6 (78.0-96.0) | 99.77% (<0.001) |
| Region of the World |  |  |  |
| Americas | 2 | 82.3 (81.9-82.6) | - |
| African | 4 | 57.7 (47.3-67.8) | 99.35% (<0.001) |
| Eastern Mediterranean | 1 | 95.0 (94.0-95.9) | - |
| Europe | 5 | 73.5 (51.5-90.7) | 99.97% (<0.001) |
| South-East Asia | 0 | - | - |
| Western Pacific | 7 | 67.7 (46.1-85.9) | 99.92% (<0.001) |

ES = Effect size; 95% CI = 95% confidence intervals

**Figure 2.**
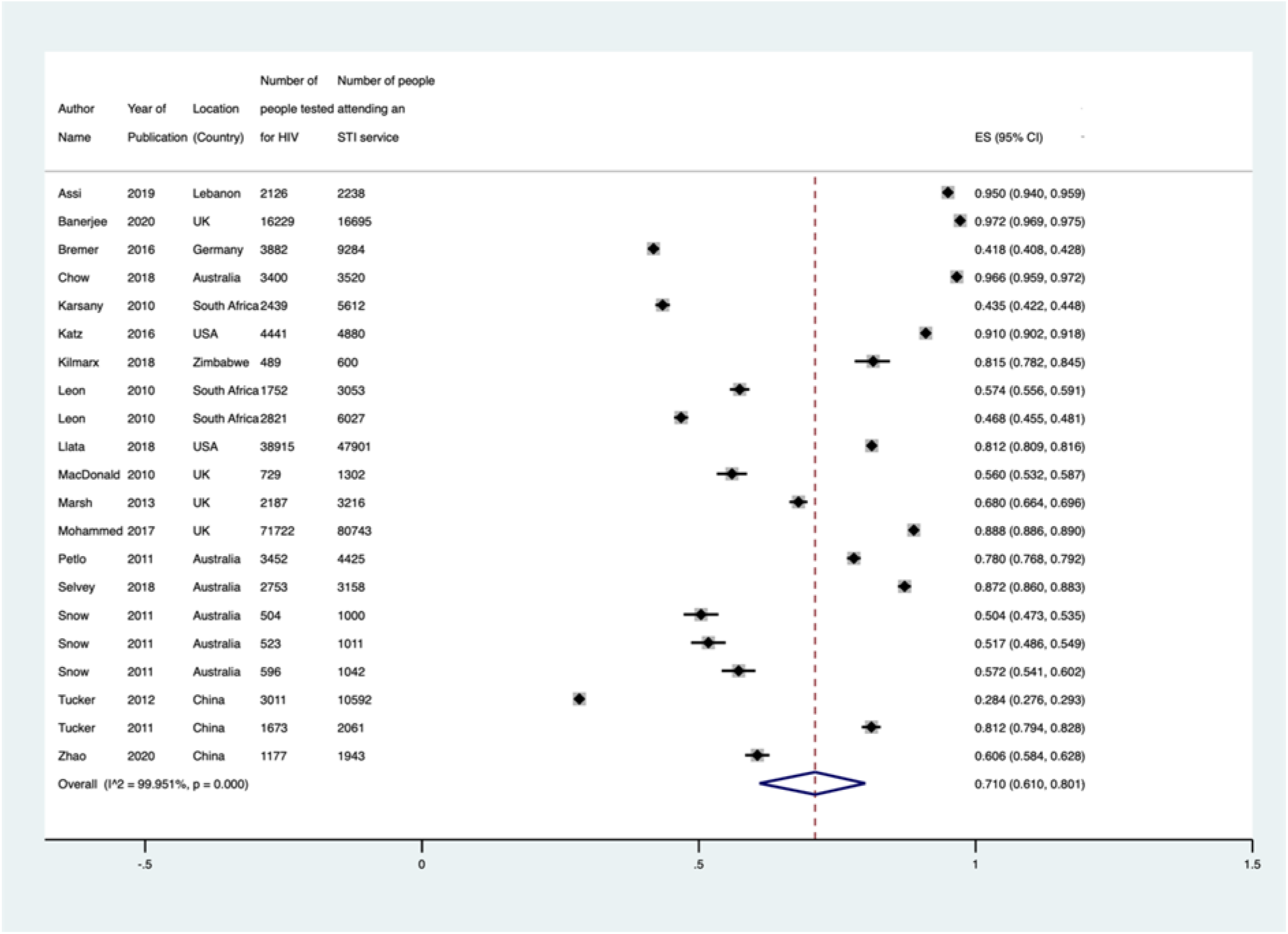
Forest plot for HIV testing among people attending a clinic with STI testing services. ES = Effect size; 95% CI = 95% confidence intervals

### Percentage of people tested for HIV among those tested for an STI

Fifteen studies provided 23 estimates from 15 studies for the meta-analysis (Figure 3, Table 3). The pooled percentage of people tested for HIV among those tested for STIs independent of the type of service was 61.3% (95% CI: 53.9-68.4%, *I*^2^=99.96%). There was evidence of publication bias (p=0.032) (Supplementary Figure 2) with a pooled prevalence of 57.8% (95% CI: 44.8-70.8) when we imputed potentially missing studies. The meta-regression analysis revealed that studies from middle-income countries compared with high-income countries (AOR 2.14, 95%CI:1.44-3.17) and regions of the world (Europe: AOR 2.04 (95%CI: 1.22-3.42); Western Pacific: AOR 1.60 (95%CI:1.04-2.44) compared with the region of the Americas, explained most of the heterogeneity (adjusted R^2^=70.5%) (Supplementary Table 3).

**Table 3.** Pooled percentage of people tested for HIV who were tested for an STI.

| | Number<br>of studies | Pooled % of people tested<br>for HIV | $I^2$ (p value) |
| --- | --- | --- | --- |
| Total | 15 | 61.3 (53.9-68.4) | 99.96% (<0.001) |
| Country income level |  |  |  |
| High | 11 | 51.1 (43.6-58.4) | 99.96% (<0.001) |
| Middle | 4 | 88.6 (85.8-91.2) | 94.44% (<0.001) |
| Type of HIV testing |  |  |  |
| Rapid HIV testing | 2 | 91.6 (89.7-93.4) | - |
| Venepuncture | 8 | 59.3 (43.4-74.3) | 98.98% (<0.001) |
| Unclear | 5 | 57.0 (44.3-69.2) | 99.76% (<0.001) |
| Recruitment site |  |  |  |
| STI clinic | 5 | 75.0 (71.2-78.7) | 95.61% (<0.001) |
| Hospital outpatients | 1 | 65.0 (61.7-68.2) | - |
| Emergency department | 4 | 18.5 (8.7-30.9) | 99.70% (<0.001) |
| General Practice | 1 | 65.0 (61.7-68.2) | - |
| Online | 1 | 100 (76.8-100) | - |
| Study population |  |  |  |
| Sexual minorities | 2 | 98.4 (93.0-100) | 53.40% (<0.001) |
| Not sexual minorities | 13 | 52.6 (44.6-60.6) | 99.97% (<0.001) |
| Latest year of study |  |  |  |
| Before 2010 | 5 | 59.8 (32.6-84.0) | 99.94% (<0.001) |
| 2010-2014 | 4 | 79.1 (60.0-93.4) | 99.82% (<0.001) |
| 2015 onwards | 6 | 48.4 (38.3-58.5) | 99.98% (<0.001) |
| Region of the World |  |  |  |
| Americas | 9 | 42.5 (33.6-51.5) | 99.97% (<0.001) |
| African | 1 | 90.1 (88.1-91.9) | - |
| Eastern Mediterranean | 0 | - | - |
| Europe | 2 | 97.9 (86.9-100) | - |
| South-East Asia | 0 | - | - |
| Western Pacific | 3 | 75.7 (71.6-79.6) | 96.18% (<0.001) |

ES = Effect size; 95% CI = 95% confidence intervals

**Figure 3.**
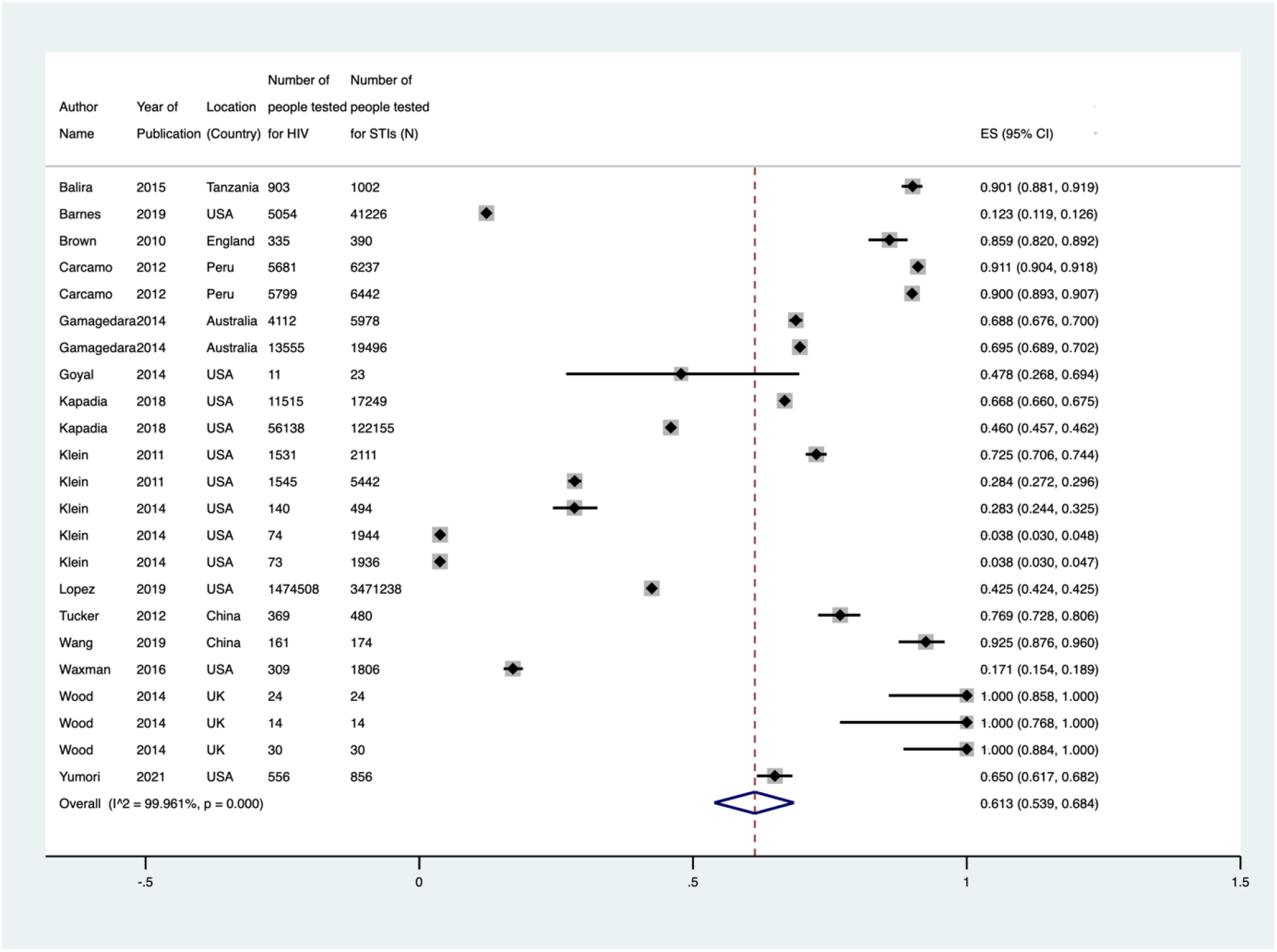
Forest plot for HIV testing among people tested for STIs. ES = Effect size; 95% CI = 95% confidence intervals

### Percentage of people tested for HIV among those diagnosed with STIs

Thirteen studies provided 22 estimates from 13 studies for the meta-analysis (Figure 4, Table 4). The pooled percentage of people tested for HIV among those diagnosed with an STI was 35.3% (95% CI: 27.1-43.9, *I*^2^=99.89%). There was no evidence of publication bias (p=0.088, Supplementary Figure 3). We could only run the univariable analysis for the meta-regression analysis as there were insufficient observations for the multivariable analysis. We found that sexual minorities (OR 1.62, 95%CI:1.16-2.25) compared with non-sexual minorities were more likely to be tested for HIV among those diagnosed with STIs (adjusted R^2^=32.5%) (Supplementary Table 4).

**Table 4.** Pooled percentage of people tested for HIV among those diagnosed with STIs.

| | Number<br>of<br>studies* | Pooled % of people tested<br>for HIV | $I^2$ (p value) |
| --- | --- | --- | --- |
| Total | 13 | 35.3 (27.1-43.9) | 99.89% (<0.001) |
| Country income level |  |  |  |
| High | 11 | 35.6 (27.1-44.6) | 99.89% (<0.001) |
| Mixed | 2 | 51.3 (49.9-52.6) | - |
| Type of HIV testing |  |  |  |
| Rapid HIV testing | 1 | 39.0 (35.9-42.2) | - |
| Venepuncture | 2 | 36.7 (29.2-44.5) | 98.94% (<0.001) |
| Unclear | 10 | 34.6 (24.3-45.7) | 99.92% (<0.001) |
| Recruitment site |  |  |  |
| STI clinic | 2 | 50.5 (49.2-51.8) | - |
| Hospital outpatients | 1 | 39.0 (35.9-42.2) | - |
| General Practice | 4 | 40.7 (34.6-47.0) | 97.77% (<0.001) |
| Study population |  |  |  |
| Sexual minorities | 1 | 81.0 (79.9-82.0) | - |
| Not sexual minorities | 12 | 32.7 (26.2-39.7) | 99.82% (<0.001) |
| Latest study year |  |  |  |
| Before 2010 | 2 | 23.5 (23.3-23.7) | - |
| 2010-2014 | 9 | 37.8 (26.9-49.4) | 99.80% (<0.001) |
| 2015 onwards | 3 | 25.4 (2.8-60.1) | - |
| Region of the World |  |  |  |
| Americas | 7 | 46.7 (29.9-63.8) | 99.96% (<0.001) |
| African | 1 | 13.4 (8.7-19.4) | - |
| Eastern Mediterranean | 0 | - | - |
| Europe | 3 | 26.7 (18.3-36.0) | 98.44% (<0.001) |
| South-East Asia | 0 | - | - |
| Western Pacific | 1 | 32.3 (29.4-35.4) | - |

\* Some categories may not add up to the total number of studies because of missing data

**Figure 4.**
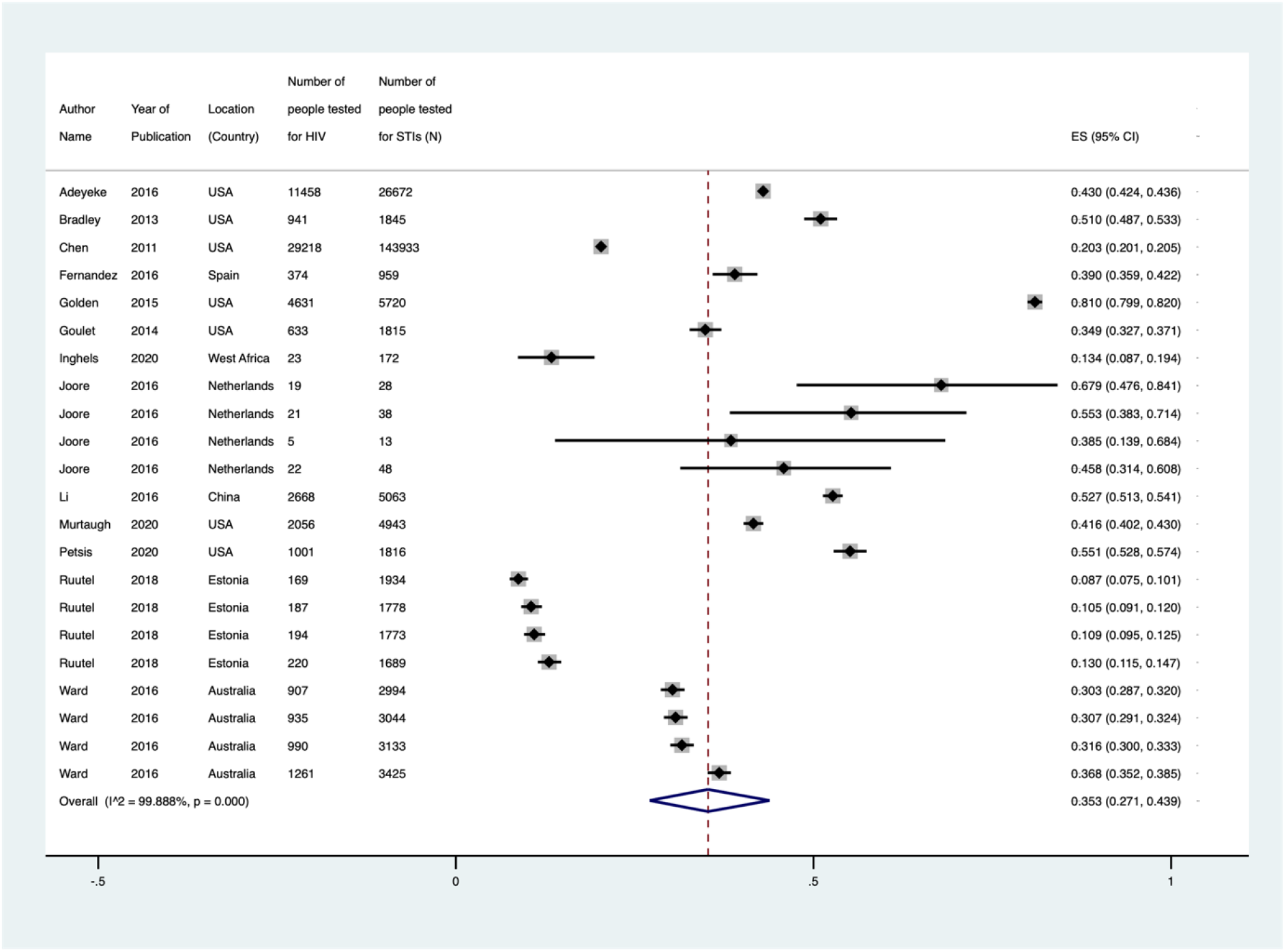
Forest plot for HIV testing among those diagnosed with an STI. ES = Effect size; 95% CI = 95% confidence intervals

### People with STI symptoms

Only three papers provided estimates for HIV testing among people presenting with STI symptoms (Figure 5). The pooled percentage was 27.1% (20.5-34.3), *I*^*2*^=91.16 (p=0.017). There was no evidence of publication bias (p=0.269, Supplementary Figure 4).

**Figure 5.**
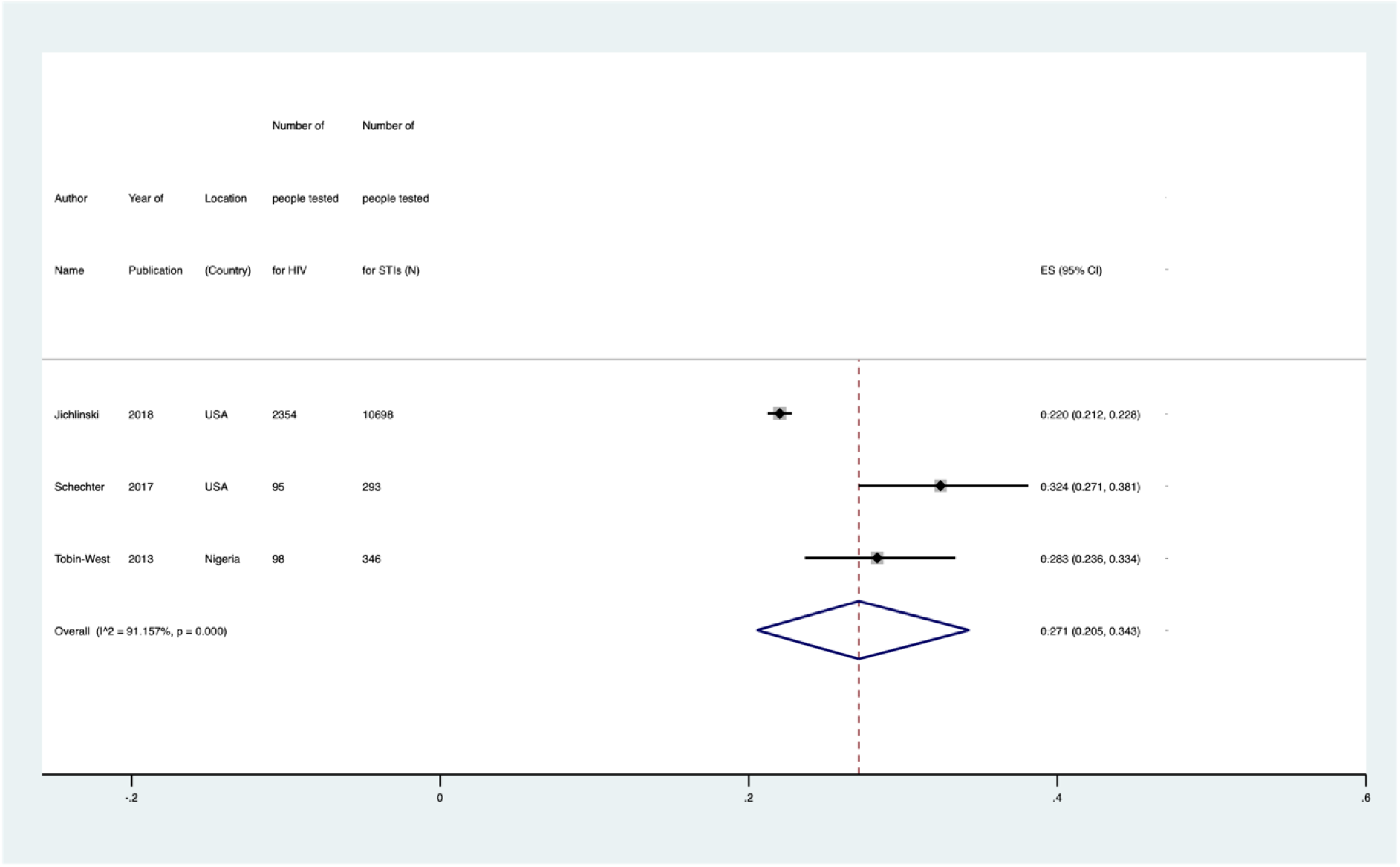
Forest plot for HIV testing among those with STI symptoms. ES = Effect size; 95% CI = 95% confidence intervals

### Factors associated with concurrent HIV/STI testing

The barriers and facilitators to concurrent HIV/STI testing can be broadly classified into individual and service factors. At an individual level, attitudes or perceptions, fear and knowledge were attributes identified. On a service level, these included the provision of services, ease of access, stigmatising features, privacy and confidentiality, and bureaucracy. Further details are provided in the Appendix, but we summarise the key findings below.

### Individual Level Barriers (Supplementary Table 5)

Three studies of varying population types (clients with STIs, clients attending an STI clinic and people attending a genitourinary medicine clinic) elaborated that a low perceived susceptibility to HIV acted as deterrence for testing; this included people who felt that they had no or little exposure to HIV risk factors [19-21]. Amongst those tested for STIs, two studies mentioned that most participants chose not to accept an HIV test as they had previously been tested [20, 22]. Fear of HIV testing (including the fear of result disclosure, needle phobia, fear of financial costs) was a common reason for refusing the test amongst all clients [21-23]. The stigma associated with HIV testing among clients attending a Nigerian STI service or a Dutch GP clinic was accompanied by the refusal and provision of HIV testing, although this was untrue among clients attending an STI Clinic in Urban China [19, 23, 24]. For clients attending an STI service or being tested for STIs, insufficient knowledge around HIV testing (unaware of testing methods and where testing can be performed) was another commonly cited reason for refusing HIV testing [25, 26]. Overall, independent of whether clients were clients with STIs or attending an STI service/GP Clinic, the individual barriers to testing were similar.

### Individual Level Facilitators

Amongst clients tested for HIV while attending an STI service, integration of HIV counselling and education (e.g. peer-based education targeting youth, provider-initiated testing and counselling (PITC)) into HIV testing was associated with increased HIV testing uptake [20, 27].

### Service Level Barriers

In a Dutch study by Joore et al.,[23] a reason for not conducting concurrent HIV testing was the concern among health providers that clients would not be able to afford additional HIV testing when presenting for STI testing. Other forms of deterrence from offering HIV testing to clients presenting for STI testing, as reported by health providers, included insufficient time during consultations, low perceived HIV risk by the clinician, and having yet to establish a relationship with new clients [23]. In some testing sites in the USA, clients presenting for STI testing were not offered HIV testing simply because of the lack of the offer to test for both HIV and STI in the same visit [28].

### Service Level Facilitators

Service factors that improved ease of access to HIV testing amongst clients who were tested for STI included implementation of dual HIV and syphilis testing[29], express testing services for lower-risk individuals [30] and convenient testing locations [26]. Internet-based HIV and STI testing, either through self-collection or allowing clients to present to designated specimen collection sites, integrated with existing clinic-based services, can increase HIV testing rates [31]. Clients being tested or diagnosed with STIs at both GP and STI clinics also felt that routine offer of HIV testing would greatly increase testing uptake [26, 32]. Raising awareness of sexual health in a non-judgemental and professional manner while maintaining confidentiality was reported to increase trust in the healthcare professional and improve the acceptability of HIV testing [26]. Additionally, national policies recommending concurrent HIV testing with STI testing can be effective amongst female sex workers in Uganda [33].

Where available, we summarised the HIV positivity, subpopulation, and recruitment site for each study population in Supplementary Tables 6-9. The risk of bias assessments is provided in the Appendix (pp33-52).

This systematic review and meta-analysis summarised the percentage of HIV testing among those who attended an STI service (71.0%), tested for STIs (61.3%), diagnosed with an STI (35.3%) or had symptoms of an STI (27.1%). To our best knowledge, this is the first attempt to collate this data to highlight the current missed opportunities for HIV testing among those already engaged in care and potentially at higher risk of HIV. Thus, strengthening strategies to improve HIV testing in these settings could help reach the UNAIDS target of diagnosing 95% of people living with HIV. Strengthening the integration of HIV and STI testing through health services is not only important for targeting HIV testing and increasing efficiencies but also for achieving broader goals within the WHO global health sector strategy[34] and the sustainable development goals to eliminate communicable diseases by 2030 [35].

Improving access points to testing could decrease the HIV testing gap. As a minimum, people who are tested for other STIs (especially if they have STI symptoms or an STI diagnosis) should be offered HIV testing, and vice versa, as the risk factors for STIs and HIV often overlap. Our study found that the testing location was a major driver of concurrent HIV/STI testing. Whilst concurrent HIV/STI testing was relatively high in STI clinics (75.0%), hospital outpatient clinics (65%) and general practice (65%), the opposite was found in emergency departments (18.5%). This lower rate of testing in emergency departments is consistent with a systematic review of HIV testing in low-resource settings, suggesting missed opportunities for better integration of HIV testing into emergency departments [36]. This could include routinely offering HIV testing to all clients being tested for other STIs (opt-out)[37, 38], improving access to HIV/syphilis dual testing or multiplex HIV/STI testing platforms, ensuring robust systems for follow-up, and providing education and training to the health workforce in line with the WHO and national testing guidelines.

This review found poor concurrent HIV/STI testing among those already diagnosed with an STI (35.3%). This is despite the WHO recommendation for the routine offer of HIV testing since 2007 (provider-initiated HIV testing) as a standard component of medical care for clients attending health facilities in high HIV burden settings and for all people with STIs in all settings [39] and for testing and retesting for people presenting with a diagnosis or receiving treatment for STIs [8]. A positive STI test is a marker of risk, and clients may often be diagnosed with multiple STIs in the same visit, particularly among men who have sex with men (MSM) and transgender populations [39-41]. An example of low concurrent HIV/STI testing among clients with STIs was in the context of HIV indicator condition-guided HIV testing in Estonia [42]. HIV indicator condition-guided HIV testing (which includes STIs as an indicator) could be an effective approach to identifying undiagnosed HIV, but there is poor awareness and adherence to this by clinical providers, limiting its impact [43]. Similarly, a study from Cote d’Ivoire reported low offers of HIV testing from medical staff for clients diagnosed with STIs. This further highlights that in many settings, health providers do not routinely offer HIV testing (even amongst those with STI symptoms). Health care worker training, standard operating procedures, and resourcing are critical to support the routine offer of HIV testing amongst those diagnosed with an STI or presenting with STI symptoms.

Among the included studies, there were several situations where concurrent HIV/STI testing was high. First, it was high in studies using the routine offer of HIV testing amongst STI clients [25, 44-46]. Routine offer of HIV testing in antenatal settings has been implemented successfully in many countries for more than a decade [47, 48]. In South Africa, the proportion of new STI clients being tested for HIV significantly increased from 42.6% to 56.4% following the universal routine offer of testing [45]. Second, we found settings that had rapid point-of-care HIV testing available markedly increased concurrent HIV/STI testing. Rapid testing for HIV/syphilis has high acceptability amongst clients[49-53] and could decrease anxiety related to waiting for results, increase convenience, and provide greater confidentiality [49, 52]. Third, when HIV testing was integrated into standard STI care protocols, this delivered more consistent performance across clinics [45]. In China, uptake of routine offer of the dual HIV/syphilis rapid testing was significantly higher when compared to isolated HIV testing at STI clinics and voluntary counselling and testing clinics [46]. In the USA, the uptake of HIV testing at the time of STI diagnosis/treatment among MSM with bacterial STIs was significantly increased from 62% to 76% following an intervention where all MSM diagnosed with STIs and their partners were offered HIV testing [54]. Fourth, involving nurses in conducting HIV tests and providing HIV chronic disease care and education could increase HIV testing uptake among STI clients [44, 45, 55]. In Australia, HIV testing rates amongst HIV negative MSM significantly increased from 41% to 47% after an STI nurse was introduced into general practice clinics [56]. The authors hypothesised that medical doctors were more willing to initiate HIV testing when nurses were able to share tasks of collecting samples and performing tests, and that nurses could spend more time with clients and thus were more likely to adhere to testing guidelines [56]. This practise is already commonplace outside high-income countries with many HIV and STI services already being fully nurse-led and in community settings where trained lay providers often conduct HIV testing.

There was limited information regarding the cost-effectiveness of concurrent HIV/STI testing beyond dual HIV/syphilis testing among antenatal populations where there is evidence of its cost-effectiveness [57]. An economic evaluation of universal HIV screening in STI clinics in the US reported that identifying clients with HIV in STI clinics was more cost-effective and could even be cost-saving compared with identifying clients with HIV in hospital inpatients [58]. People with HIV who attended STI clinics were more likely to have higher CD4 counts at the time of diagnosis, allowing for earlier ART initiation [58]. In terms of staff resources, a study in South Africa reported that it was efficient for STI nurses to integrate HIV screening into their consultations [32]. This shift in responsibilities of STI nurses was achieved with relatively short training and by slightly extending their consultation time. A nurse-led express ‘Test and Go’ HIV/STI testing service for MSM in Melbourne also effectively reduced consultation costs of seeing these men [59]. A modelling analysis of implementing HIV/syphilis dual testing among key populations in Vietnam reported that annual or biannual dual testing could be cost-effective [60]. Further studies of the cost-effectiveness of integrating HIV screening with STI testing in a range of settings, especially in LMICs, would be helpful to support decision making.

The strength of this study is that we systematically reviewed the literature to synthesise knowledge on concurrent HIV/STI testing across a range of settings. This highlighted missed opportunities for HIV testing among individuals at higher risk of infection, specifically those with STI symptoms or an STI diagnosis. Our study had some limitations. First, we only included published data, and most were from a HIC setting, especially from the US. Therefore, our findings may not be generalisable to LMICs or in settings with a high HIV burden. Second, we found significant heterogeneity in our meta-analysis not explained by sampling variability alone. This heterogeneity may include unmeasured confounders between studies related to patient population characteristics (e.g., background HIV risk, distribution of socioeconomic status), recruitment methods, and provider-level factors (e.g., perception of the need to test clients, time and cost constraints). Nevertheless, our study findings highlight high proportions of missed opportunities to test for HIV. Finally, this review of published literature, although indicating current practices and gaps, may not reflect broad practice and more work is needed to assess the programme implementation landscape. Published studies may prioritise services that are currently understating some level of HIV testing and/or efforts to increase or improve efficiencies and there may be even greater gaps and missed opportunities, including in LMIC. Data from the Global AIDS Monitoring (GAM) reports that 16% of reporting countries 31/194 in 2021 had a policy of offering dual HIV/syphilis testing for key populations. However, the extent to which this is implemented is not reported.

## Conclusions

In conclusion, we identified significant gaps in concurrent HIV/STI testing globally, specifically amongst people diagnosed with an STI. We suggest better integration of HIV and STI services, particularly routinely offering HIV testing to all people with STI diagnosis and symptoms, to increase HIV diagnosis in this population at higher HIV risk.

## Supporting information

Additional File 3- Search Strategy

Additional File 4- Quality Checklists

## Data Availability

All data produced in the present work are contained in the manuscript.

## Authors’ contributions

JJO designed the research study. JJO, KS, ET and AL performed the research and analysed the data. KS, ET, AL, ML, RB, MBM, MSJ, MBD, CJ, SLG, EC, CKF, JJO wrote the paper.

## Author information

The findings and conclusions in this paper are those of the authors and do not necessarily represent the official position of the World Health Organization.

## Acknowledgements

Not applicable.

## Additional Files

### Additional file 1: Supplementary Figures

- Supplementary Figure 1 Funnel plot for HIV testing among people attending an STI service
- Supplementary Figure 2 HIV testing among people tested for STIs
- Supplementary Figure 3 Funnel plot of those diagnosed with an STI
- Supplementary Figure 4 Funnel plot for people with STI symptoms
- Supplementary Figure 5 World map of included studies

### Additional file 2: Supplementary Tables

- Supplementary Table 1 Characteristics of included studies
- Supplementary Table 2 Meta-regression results for HIV testing for those attending a clinic with STI testing services
- Supplementary Table 3 Meta-regression results for HIV testing for those tested for an STI
- Supplementary Table 4 Meta-regression results for HIV testing for those diagnosed with an STI
- Supplementary Table 5 Factors associated with concurrent HIV and STI testing amongst STI patients
- Supplementary Table 6 HIV positivity among people tested for HIV who attended an STI service
- Supplementary Table 7 HIV positivity among people who were tested for STIs
- Supplementary Table 8 HIV positivity among people who were tested for STIs
- Supplementary Table 9 HIV positivity among people with STI symptoms

### Additional file 3: Search Strategy

Information detailing the search strategy used for the systematic review.

### Additional file 4: Quality Checklists

Tables detailing the quality scores using the Cochrane risk of bias tool for randomised controlled trials, Newcastle-Ottawa quality assessment scale for cross-sectional, cohort and case-control studies, consolidated health economic evaluation reporting standards (CHEERS) checklist for economic evaluation studies, and Joanna Briggs Institute (JBI) critical appraisal checklist for qualitative studies.

