## Additional file 1: Supplementary Figures for "Missed opportunities for HIV testing among those who accessed sexually transmitted infection (STI) services, tested for STIs and diagnosed with STIs: a systematic review and meta-analysis"

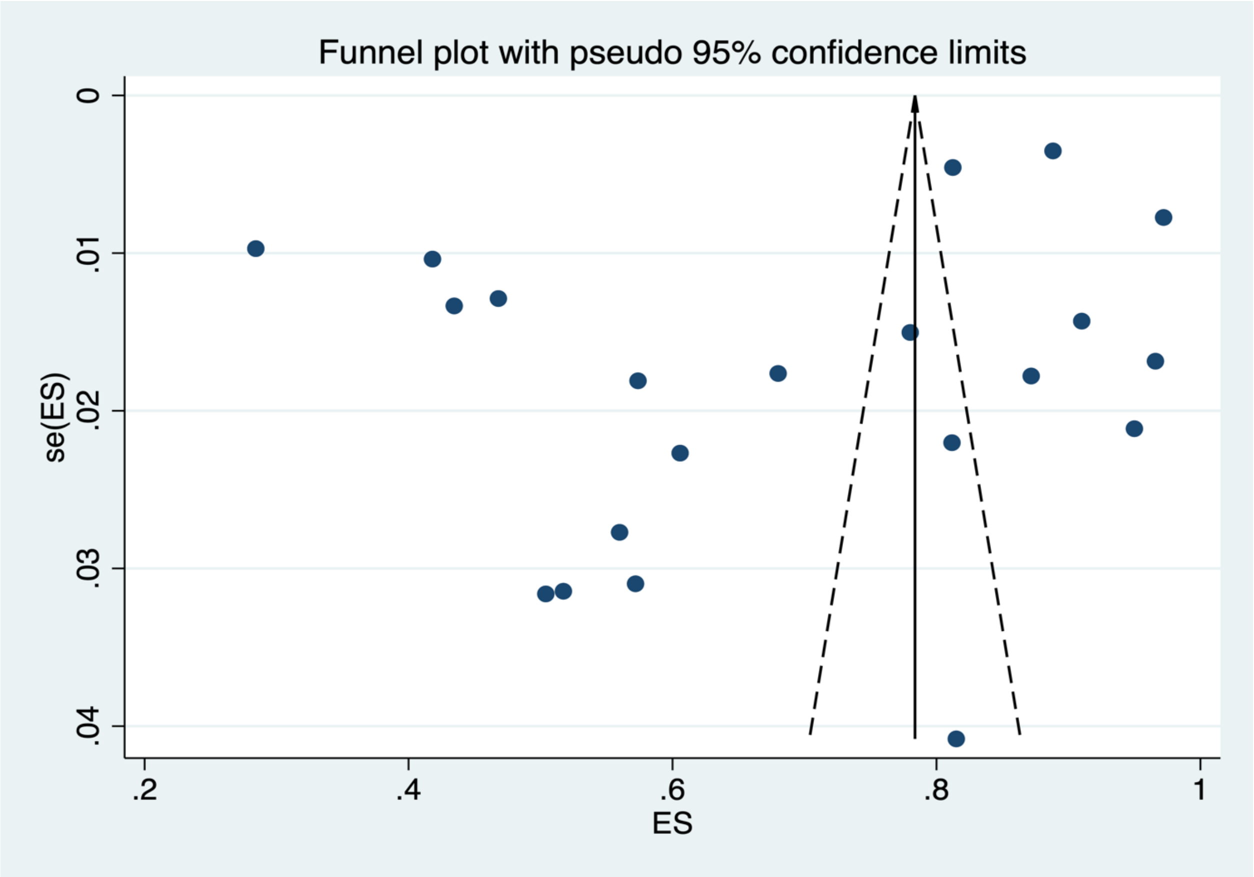


**Supplementary Figure 1 Funnel plot for HIV testing among people attending an STI service**

**Egger's test, p=0.837**

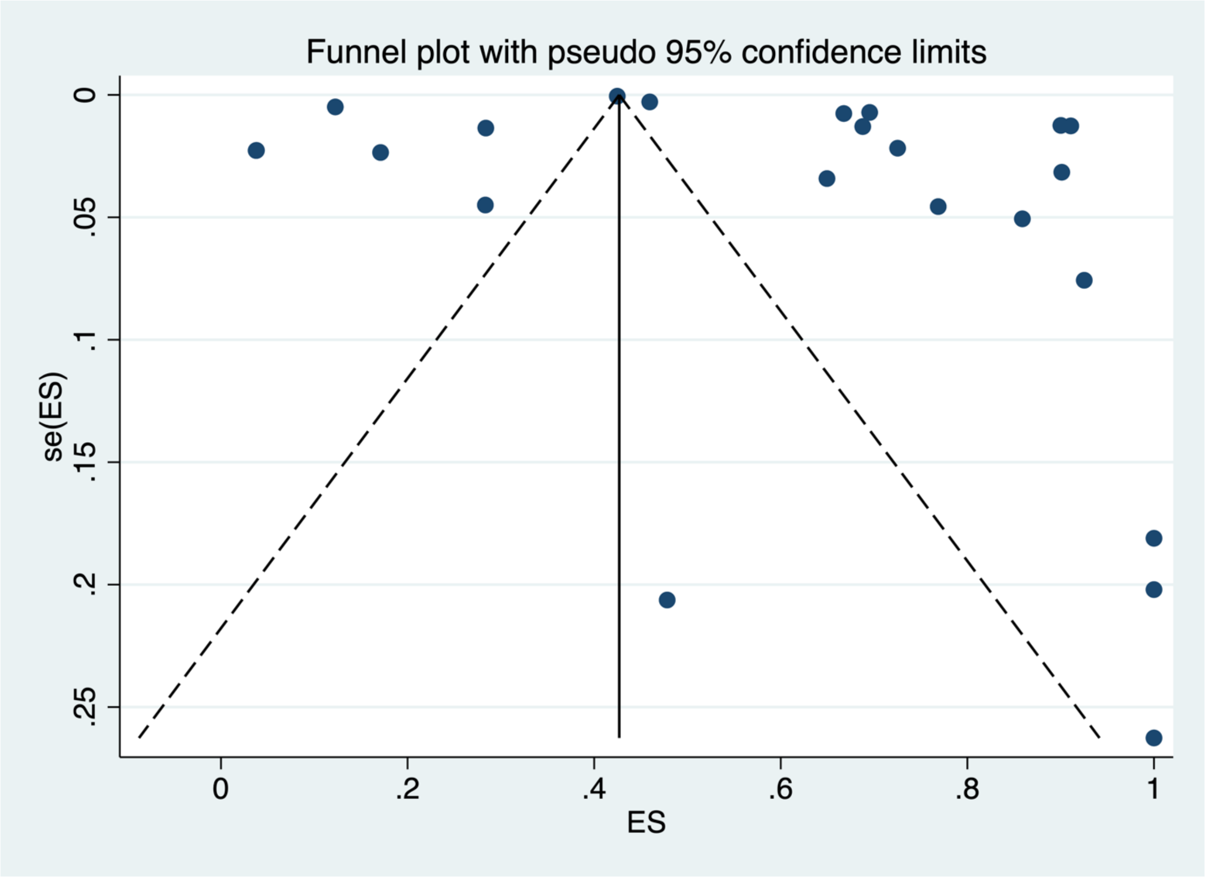


**Supplementary Figure 2 HIV testing among people tested for STIs**

**Egger's test 0.032**

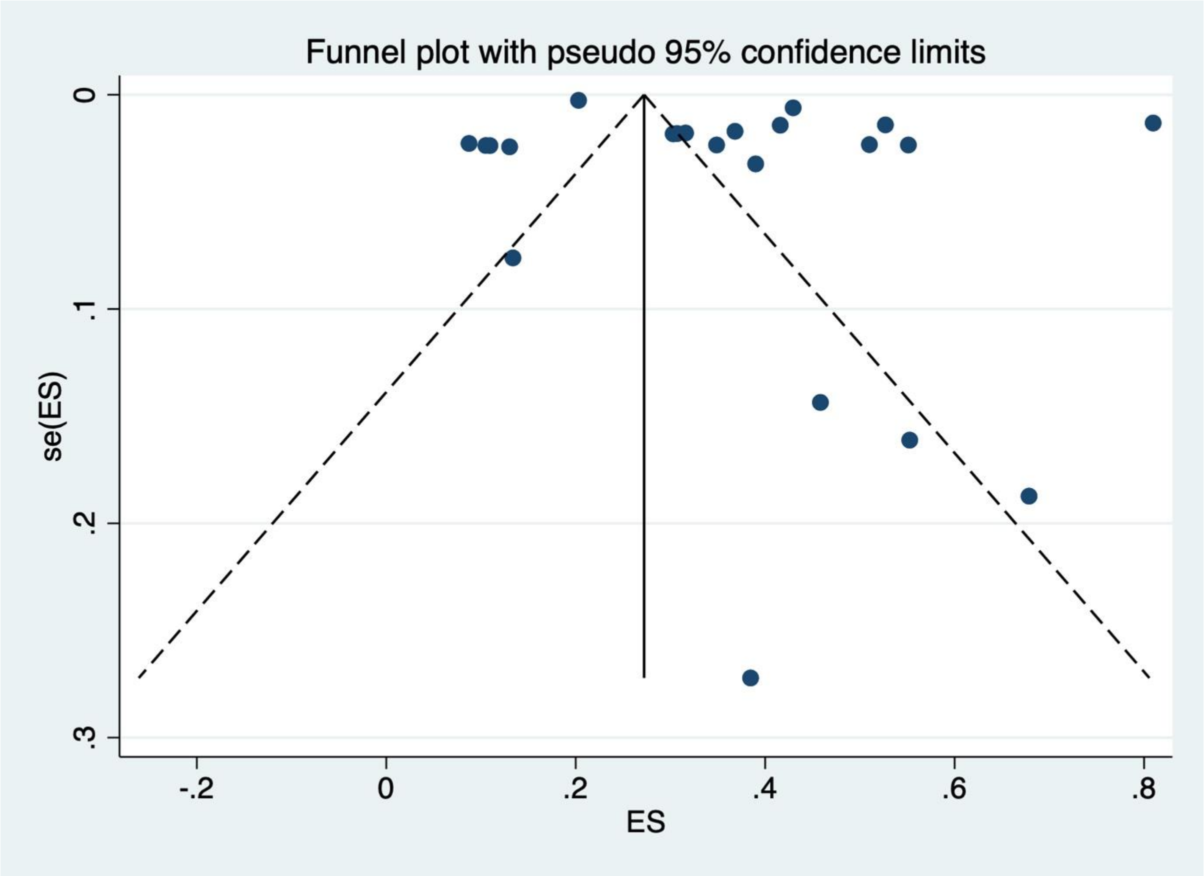


**Supplementary Figure 3 Funnel plot of those diagnosed with an STI**

**Egger's test, p=0.088**

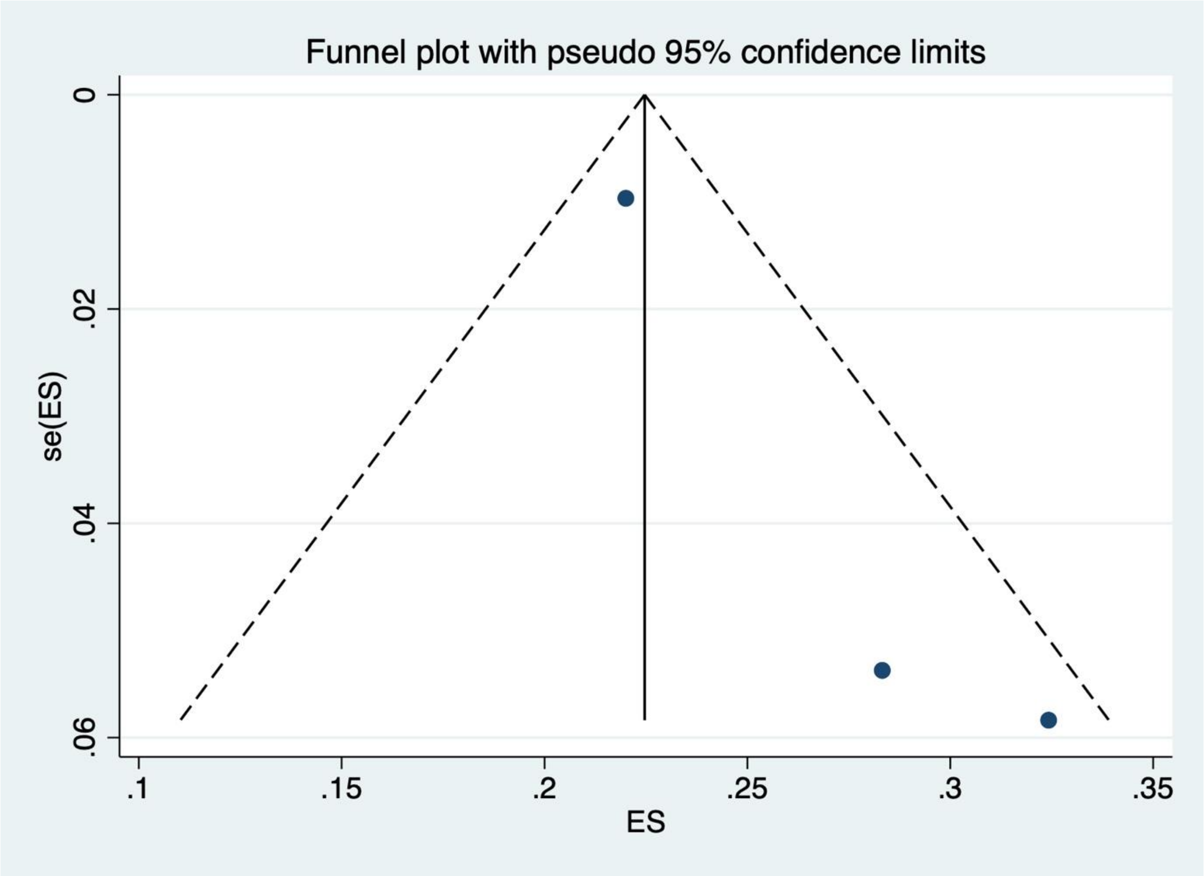


**Supplementary Figure 4 Funnel plot for people with STI symptoms**

**Egger p=0.269**

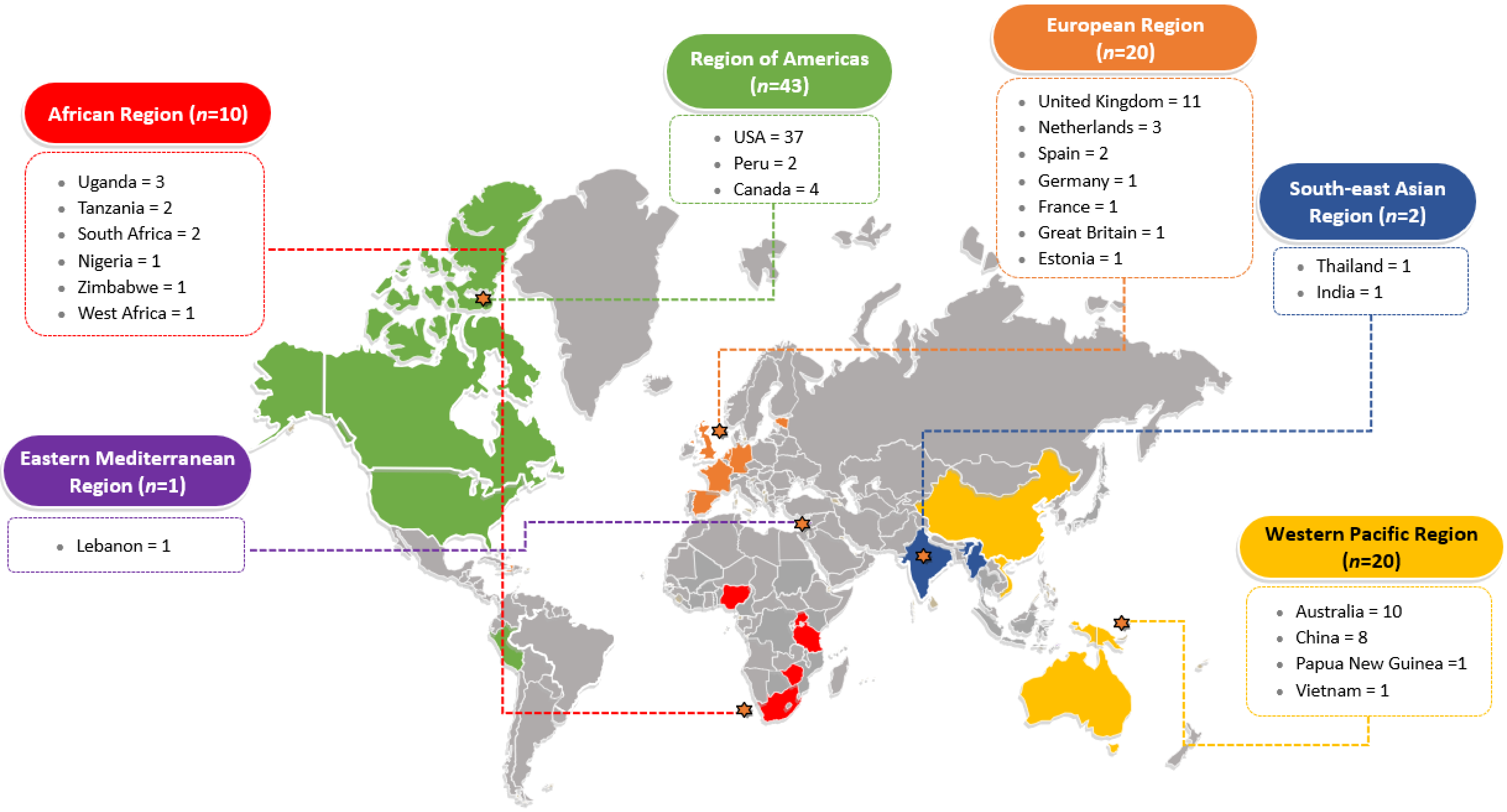


**Supplementary Figure 5 World map of included studies**
