## Additional File 2- Supplementary Tables for "Missed opportunities for HIV testing among those who accessed sexually transmitted infection (STI) services, tested for STIs and diagnosed with STIs: a systematic review and meta-analysis"

**Supplementary Table 1 Characteristics of included studies**

| Author Name | Year of study | Type of Study | Location (Country) | Study Population |
|---|---|---|---|---|
| Adam[1] | 2011 | Observational | Australia | Sexual minorities |
| Adekeye[2] | 2009 | Observational | USA | General |
| Assi[3] | 2015-2018 | Observational | Lebanon | Sexual minorities |
| Avoundjian[4] | 2014-2016 | Observational | USA | Sexual minorities |
| Badman[5] | 2014 | Observational | Papua New Guinea | Pregnant women |
| Baker U[6] | 2011-2014 | Observational, Qualitative | Tanzania, Uganda | Pregnant women |
| Balan C [7] | 2016-2017 | Qualitative | USA | Sexual minorities |
| Balira[8] | 2008-2009 | Observational | Tanzania | Healthcare workers & Pregnant women |
| Banerjee[9] | 2017 | Observational | UK | General |
| Barber[10] | | Observational | Australia | Sexual minorities |
| Barnes[11] | 2014-2015 | Observational | USA | General |
| Bauermeister[12] | 2013 | Observational | USA | Sexual minorities |
| Bauermeister[13] | | Experimental | USA | Sexual minorities |
| Beck[14] | | Modelling | USA | Sexual minorities, Youth |
| Bien[15] | | Qualitative | China | Sexual minorities |
| Bradley H[16] | 2009-2010 | Observational, Qualitative | USA | General |
| Bremer[17] | 2010-2011 | Observational | Germany | Sex workers |
| Bristow[18] | | Observational | Peru | Sexual minorities |

| | | | | |
|---|---|---|---|---|
| Brown L[19] | 2006 | Experimental | UK | General |
| Carcamo[20] | 2002 | Observational | Peru | General |
| Cayuelas [21] | 2013 | Observational | Spain | General |
| Chen J[22] | 2007 | Observational | USA | General |
| Chow E[23] | 2015-2016 | Observational | Australia | Sexual minorities |
| Cushman[24] | 2014 | Observational | USA | Sexual minorities |
| Fernandez[25] | 2008-2010 | Observational | Spain | General |
| Gamagedara[26] | 2009 | Observational | Australia | General |
| Gilbert, M.[27] | 2015-2016 | Observational | Canada | General |
| Golden[28] | 2010-2014 | Observational | USA | Sexual minorities |
| Goulet[29] | 2010 | Observational | USA | Veterans |
| Goyal[30] | 2011 | Observational | USA | Youth |
| Goyal, M. K.[31] | 2011 | Observational | USA | Youth |
| Heard[32] | | Observational | Australia | Sexual minorities |
| Hottes[33] | 2011 | Qualitative | Canada | Sexual minorities |
| Inghels[34] | 2017 | Observational | West Africa | General |
| Jichlinski[35] | 2010 to 2015 | Observational | USA | Youth |
| Jones[36] | 2013 | Qualitative | UK | Youth |
| Joore[37] | 2014 | Qualitative | Netherlands | General |
| Joore[38] | 2008-2013 | Observational | Netherlands | General |
| Joore[39] | 2009-2013 | Observational | Netherlands | General |
| Josten[40] | 2017 | Observational | USA | Youth providers |
| Kapadia[41] | 2010-2015 | Observational | USA | General |
| Kharsany[42] | 2005-2006 | Observational | South Africa | Women attending STI clinics |
| Katz[43] | 2012-2014 | Observational | USA | Sexual minorities |
| Kilmarx[44] | 2014-2015 | Observational | Zimbabwe | General |
| Klein[45] | 2009-2011 | Observational | USA | General |

| | | | | |
|---|---|---|---|---|
| Klein[46] | 2009 | Observational | USA | General |
| Knight[47] | | Qualitative | Canada | Sexual minorities |
| Lanier[48] | 2010-2011 | Observational, Qualitative | USA | Physicians |
| Leon[49] | 2007 | Pragmatic cluster non-randomised controlled trial | South Africa | General |
| Li[50] | 2013-2014 | Observational | | General |
| Llata[51] | 2010-2013 | Observational | USA | Sexual minorities |
| Lopez[52] | 2010-2015 | Observational | USA | General |
| MacDonald[53] | 2004 | Observational | UK | General |
| Marsh[54] | 2011 | Observational | UK | Prisoners |
| Maxwell[55] | | Observational | UK | General Practitioners |
| McDonagh[56] | | Qualitative | UK | Sexual minorities |
| Mohammed [57] | 2014 | Observational | UK | Black Africans attending sexual health clinics |
| Moore[58] | 2014 | Observational | USA | Youth |
| Moore[59] | 2010-2011 | Observational | USA | Youth |
| Muhindo [60] | 2018 | Observational | Uganda | Sex workers |
| Muhindo [61] | 2019 | Observational | Uganda | Sex workers |
| Mullens[62] | 2016-2017 | Observational, Qualitative | Australia | Sexual minorities |
| Murtaugh[63] | 2013 | Observational | USA | General |
| Ngo[64] | 2007-2009 | Observational | Vietnam | Youth |
| Owusu-Edusei Jr[65] | 2012 | Observational | USA | Sexual minorities |
| Owusu-Edusei Jr[66] | | Modelling | China | Pregnant women |

| | | | | |
|---|---|---|---|---|
| Pai[67] | 2008-2009 | Observational | India | Pregnant women |
| Petlo[68] | 2003-2009 | Observational | Australia | Sexual minorities |
| Petsis[69] | 2014-2017 | Observational | USA | Youth |
| Phrasisombath[70] | 2010 | Qualitative | Thailand | Sex workers |
| Prabhu[71] | 2006-2008 | Modelling | USA | General<br>Sex workers |
| Rocchetti[72] | 2013 | Observational | France | General Practitioners |
| Ruutel[73] | 2012 | Observational | Estonia | People with indicator conditions |
| Saunders[74] | 2010 | Observational | Great Britain | Youth |
| Schechter[75] | 2011-2015 | Observational | USA | General |
| Scheim[76] | 2013 | Qualitative | Canada | Sexual minorities |
| Selvey[77] | 2011-2015 | Observational | Australia | Sexual minorities |
| Sharma[78] | 2017-2018 | Observational | USA | Sexual minorities,<br>Youth |
| Slinkard[79] | | Qualitative | USA | Older adults |
| Snow[80] | 2006-2007 | Observational | Australia | Sexual minorities |
| Sullivan[81] | | Observational | USA | Sexual minorities |
| Tobin-West[82] | 2011 | Observational | Nigeria | General |
| Tucker[83] | 2009 | Observational | Guangdong China | Patients at STI Clinic |
| Tucker[84] | 2009 | Observational | South China | General |
| Tucker[85] | 2009-2010 | Observational | China | General |
| Underhill[86] | 2012-2014 | Qualitative | USA | Sexual minorities |
| Wang[87] | 2018 | Observational | China | Sexual minorities |
| Wang [88] | 2002 | Observational | China | General |
| Ward[89] | 2010-2014 | Observational | Australia | Aboriginal communities |
| Ward[89] | 2011 | Observational | Australia | Aboriginal communities |

| | | | | |
|---|---|---|---|---|
| Waxman[90] | 2015 | Observational | USA | General |
| Williford[91] | 2015-2019 | Observational | USA | General |
| Wood[92] | 2013 | Observational | UK | Sexual minorities |
| Youssef[93] | 2012 | Observational | UK, Brighton | General |
| Yumori[94] | 2019 | Observational | USA | General |
| Zhao[95] | 2016 | Observational | China | General |

**Supplementary Table 2 Meta-regression results for HIV testing for those attending a clinic with STI testing services**

| Characteristic | Variable | OR (95% CI) | p value | AOR (95% CI) | p value |
|---|---|---|---|---|---|
| Country income level | High | 1 | | 1 | |
| | Middle | 0.89 (0.74-1.09) | 0.24 | 0.94 (0.77-1.14) | 0.41 |
| Type of HIV testing | Rapid HIV testing | 1 | | 1 | |
| | Venepuncture | 0.89 (0.68-1.15) | 0.33 | * | |
| Study population | Not sexual minorities | 1 | | 1 | |
| | Sexual minorities | 1.15 (0.96-1.39) | 0.13 | 1.37 (1.10-1.71) | 0.02 |
| Latest year of study | Before 2010 | 1 | | 1 | |
| | 2010-2014 | 1.10 (0.89-1.36) | 0.36 | 1.68 (1.17-2.40) | 0.02 |
| | 2015 onwards | 1.28 (1.07-1.54) | 0.01 | 1.46 (1.26-1.69) | <0.01 |
| Region of the world | Americas | 1 | | 1 | |

| | | | | | |
|---|---|---|---|---|---|
| | African | 0.87 (0.68-1.10) | 0.23 | 1.27 (0.90-1.80) | 0.13 |
| | Eastern Mediterranean | 1.32 (0.85-2.05) | 0.20 | * | |
| | Europe | 1.02 (0.81-1.29) | 0.83 | * | |
| | Western Pacific | 0.95 (0.78-1.16) | 0.61 | 1.04 (0.79-1.37) | 0.73 |

Adjusted R-squared for multivariable model = 91.3%
* omitted because of collinearity
AOR = adjusted odds ratio; OR = odds ratio; 95% CI = 95% confidence intervals

**Supplementary Table 3 Meta-regression results for HIV testing for those tested for an STI**

| Characteristic | Variable | OR (95% CI) | p value | AOR (95% CI) | p value |
|---|---|---|---|---|---|
| Country income level | High | 1 | | 1 | |
| | Middle | 1.47 (1.09-1.96) | 0.01 | 2.14 (1.44-3.17) | <0.01 |
| Type of HIV testing | Rapid HIV testing | 1 | | 1 | |
| | Venepuncture | 0.69 (0.38-1.25) | 0.20 | 1.00 (0.42-2.41) | 1.00 |
| Study population | Not sexual minorities | 1 | | 1 | |
| | Sexual minorities | 1.56 (1.07-2.28) | 0.02 | 0.98 (0.45-2.15) | 0.95 |
| Latest year of study | Before 2010 | 1 | | 1 | |
| | 2010-2014 | 1.18 (0.86-1.62) | 0.30 | 1.18 (0.74-1.87) | 0.44 |

| | | | | | |
|---|---|---|---|---|---|
| | 2015 onwards | 0.85 (0.63-1.15) | 0.29 | 1.31 (0.92-1.87) | 0.11 |
| Region of the world | Americas | 1 | | 1 | |
| | African | 1.39 (0.71-2.69) | 0.32 | 1.00 (0.36-2.79) | 0.99 |
| | Europe | 1.52 (1.04-2.22) | 0.03 | 2.04 (1.22-3.42) | 0.01 |
| | Western Pacific | 1.24 (0.87-1.77) | 0.22 | 1.60 (1.04-2.44) | 0.04 |

Adjusted R-squared for multivariable model = 70.5%
AOR = adjusted odds ratio; OR = odds ratio; 95% CI = 95% confidence intervals

**Supplementary Table 4 Meta-regression results for HIV testing for those diagnosed with an STI**

| Characteristic | Variable | OR (95% CI) | p value |
|---|---|---|---|
| Country income level | High | 1 | |
| | Mixed | * | |
| Type of HIV testing | Rapid HIV testing | 1 | |
| | Venepuncture | 0.98 (0.71-1.36) | 0.86 |
| Study population | Not sexual minorities | 1 | |
| | Sexual minorities | 1.62 (1.16-2.25) | 0.01 |
| Latest year of study | Before 2010 | 1 | |
| | 2010-2014 | 1.09 (0.88-1.34) | 0.41 |

| | | | |
|---|---|---|---|
| | 2015 onwards | 0.91 (0.71-1.17) | 0.45 |
| Region of the world | Americas | 1 | |
| | African | 0.80 (0.52-1.23) | 0.29 |
| | Europe | 0.88 (0.74-1.05) | 0.14 |
| | Western Pacific | 0.97 (0.77-1.22) | 0.77 |

AOR = adjusted odds ratio; OR = odds ratio; 95% CI = 95% confidence intervals

**Supplementary Table 5 Factors associated with concurrent HIV and STI testing amongst STI patients**

Individual Level Barriers

| | | |
|---|---|---|
| Individual attitudes/perceptions | Low perceived susceptibility to HIV/STI | Tucker, 2012[83] - Research conducted in urban China population shows that low perceived HIV risk was the most common reason for not receiving a HIV test, approximately ½ of STD patients did not believe they have HIV risk factors, despite 40% reporting a previous STD and an increasing burden of HIV in the region.<br><br>Zhao, 2020 [95]- Among 1943 participants who attended an STD clinic in China, the most common reason for refusing HIV testing was because they felt that it was not necessary (71.5%).<br><br>Macdonald, 2010 [53]- A regional audit in Scotland, looking at HIV testing in genitourinary medicine clinics revealed that the top reason for declining a HIV test was that 26% of participants felt they were at low risk for HIV. |
| | Perception of being previously tested and hence felt that testing was not required | Kharsany, 2010 [42]- A study looking at uptake of provider-initiated HIV testing and counselling among women attending an urban STI clinic in South Africa, showed that the most common reason for not accepting a HIV test was patients had reported already having being tested (61.8%) and of these only 3.3% disclosed being HIV postive and 1.8% reported being on antiretroviral treatment.<br><br>Zhao, 2020 [95]- Among 1943 participants who attended an STD clinic in China, 14.6% refused HIV testing as they had received a test before |
| | Fear of testing and subsequent results | Kharsany, 2010 [42]- A study looking at uptake of provider-initiated HIV testing and counselling among women attending an urban STI clinic in South Africa, showed that 32.5% of participants were afraid to test or felt unready to test.<br><br>Joore, 2016 [37]- GPs working in urban or rural Dutch practices stated that indicator condition guided testing might create unnecessary fear among patients. |

| | | |
|---|---|---|
| | | Macdonald, 2010 [53]- A regional audit in Scotland, looking at HIV testing in genitourinary medicine clinics revealed that the reasons for declining a HIV test was that 15% of participants were needle phobic.<br>2% did not want to know the result |
| HIV and STI related stigma | Negative stigma associated with HIV and STI | Tobin, 2013 [82]- A study conducted in Nigeria on consequences of stigma and underutilisation of HIV screening showed that 229 (38.4%) of respondents had strong stigmatisation tendencies towards HIV positive persons compared to 367 (61.6%) with low stigma outlook. Most with high stigma tendencies, 164 (71.6%) had not tested for HIV themselves compared to 206 (56.1%) with low stigma attitudes. This indicates that stigma makes individuals reluctant to access HIV testing, treatment and care, thereby compromising attempts to fight the epidemic in general.<br><br>Tucker, 2012 [96]- In a study conducted in China in urban STD patients, fear of discrimination and loss of face was **infrequently** 7.7% reported as a reason to refuse HIV testing at STD clinic, suggesting that perceived HIV stigma is unlikely a major contributor to low HIV testing in public STD clinics. This might be subjected to reporting bias as the study only included self-reported HIV testing behaviour.<br><br>Joore, 2016 [37]- GPs working in urban or rural Dutch practices were worried about patients' responses to them regarding judging their patients' sexual behaviour and raising stigma if they discussed a HIV test when indicator conditions were diagnosed, stating that combining HIV test with other laboratory tests as a possible way to implement the routine offer of testing approach. |
| Knowledge | Insufficient knowledge regarding benefits of HIV testing or methods available | Mohammed, 2016 [57]- A study on the refusal of HIV testing among Black Africans in England showed that there was an increased likelihood of test refusal among those living in more deprived areas is consistent with evidence of poorer sexual health among this population sub- group. The reasons for this are unknown, but it may reflect a lack of knowledge about HIV.<br><br>Jones, 2017 [36]- In a qualitative interview study among young adults in Europe, lack of knowledge on testing methods available and where testing can be performed (at home, immediately in the practice, behind a curtain, in the toilet, or in another room); understanding 3Cs and HIV will inform their |

| | | |
|---|---|---|
| | | assessment of personal risk & increase confidence in utilitising it, which in turn may inform and change their health-seeking behaviours and sexual health risk behaviours. |

**Individual Level Facilitators**

| | | |
|---|---|---|
| Education | Sexual health education promotion | Ngo, 2013 [64]- An educational intervention on integration of HIC and sexual and reproductive health services for young people in Vietnam resulted in a significant increase in percentage of youth who wanted to obtain a HIV test 33 to 55%, who ever had a test from 7.5% to 15% and a repeat test in the last 12 months from 54.5% to 67.5%. Willingness to pay for a test increased from 54.5% to 75.5%.<br><br>Zhao, 2020 [95]- Among 1943 participants who attended an STD clinic in China, having received any kind of HIV/ STD-related knowledge in the last 12 months (aOR: 1.94, 95% CI 1.58 to 2.40) were positively associated with HIV testing uptake. |

**Service Level Barriers**

| | | |
|---|---|---|
| Costs | Cost of HIV test | Joore, 2016 [37]- A study interviewing 9 GPs working in urban or rural Dutch practices revealed that some GPs were concerned that patients would refuse the HIV test because of the cost. In the Netherlands, GP consultations are covered by mandatory health insurance but all insured persons have to pay a compulsory excess themselves, worry was specifically acute amongst participants working in multicultural and socioeconomically deprived areas. |
| Provision of services | Insufficient time | Joore, 2016 [37]- A study interviewing 9 GPs working in urban or rural Dutch practices, showed that GPs were concerned about the additional time taken to discuss a HIV test. |
| Perceived susceptibility by clinician | Low perceived susceptibility by clinical | Joore, 2016 [37]- GPs working in urban or rural Dutch practices were more likely to test for HIV when there is a clear link with immunodeficiency, as the relationship between HIV and immunodeficiency is clear. They also suggest indicator condition testing implemented in a high-prevalence area, with age restrictions - otherwise testing all patients is unnecessary due to low perceived risk. |

| | | |
|---|---|---|
| Availability of tests | Lack of availability of dual testing (HIV and STI) | Moore, 2013 [59]- Among 292 heterosexual college students at a midwestern university in USA, Chlamydia/gonorrhea testing rates were 43.5%. More than a third of the sample (35.6%) had been tested for both HIV and chlamydia/gonorrhea, which represented 66.2% of those who stated that they had been tested at all. This highlights the fact that less than half of sexually experienced students getting tested are seeking both HIV and chlamydia/gonorrhea testing. This finding may be due to their frequenting testing sites that do not offer both types of testing. |

**Service Level Facilitators**

| | | |
|---|---|---|
| Ease of Access | Dual testing | Bristow, 2018 [18]- Implementing a preferred and acceptable dual testing strategy. |
| | Express testing | Gamagedara, 2014 [26]- This study conducted in Melbourne, Australia among sexual health clinic attendees regarding an evaluation of express testing indicated a marginal increase in the number of clients being tested for HIV and chlamydia (1% for HIV and 2% for chlamydia) with the implementation of express testing. All clients offered a first-void urine test for chlamydia and an option to have blood tests for HIV and syphilis. This is attributed to the reduction in the time taken to see uncomplicated patients, three to four express clients can be seen in the time a routine consultation will take, translating to savings of A$60,000. Additionally, this express testing service would help to increase capacity, allowing high-risk clients with complications to access the service. |
| | Convenient testing locations | Jones, 2017 [36]- A qualitative interview study among young adults in Europe indicated that participants preferred convenience facilitated by proximity to the test location. |
| | Home-based testing/self-collected testing | Hottes, 2012 [33]- A study on the feasibility and acceptability of Internet-based STI testing among largely MSM participants in Canada, showed that the perceived benefits of Internet-based STI testing included anonymity, convenience, and client-centered control. Trust in the new online service, however, is a prerequisite to client uptake and may be engendered by transparency of information about the model, and by accounting for concerns related to confidentiality, data usage, and provision of positive (especially HIV) results. |

| | | |
|---|---|---|
| Routine offering of tests | Offering of tests as part of routine consultations | Jones, 2017[36] - In a qualitative interview study among young adults in Europe, many participants reported that sexual health should be raised routinely in consultations, important for staff to indicate that the offer was made as the patient met the target demographic, eliminating perception of being given offer due to suspected likelihood of having a STI.<br><br>Joore, 2016 [37]- A study interviewing 9 GPs working in urban or rural Dutch practices revealed that participating GPs felt that offering HIV tests to new patients registering at their practice was inappropriate as they have yet to establish a relationship with these individuals, commenting that testing for HIV is indirectly linked to somebody's personal lifestyle.<br><br>Leon, 2010 [49]- A study in Cape Town, South Africa, showed that streamlined provider-initiated, opt out HIV testing protocol delivered by nurses in busy primary healthcare clinics appears to be effective and feasible, Despite significantly higher proportion intervention group declining testing when offered (26.7% intervention versus 13.5% control, p = 0.0086). |
| Awareness | Raising awareness | Jones, 2017[36]- Among a qualitative interview study among young adults in Europe, 2 participants felt that awareness should be raised around GP practices offering sexual health services. |
| Privacy, confidentiality and anonymity | Presence of trust helps promote discussion of HIV testing | Jones, 2017 [36]- A qualitative interview study among young adults in Europe showed that participants preferred GPs that they trust, or those who showed professionalism when discussing sexual health. |
| Policy | Presence of policy encouraging HIV testing | Muhindo, 2019 [60]- A study on the psychosocial correlates of regular syphilis and HIV screening practices among female sex workers showed that focuse on HIV screening policies in Uganda and Kampala has resulted in higher attitudes and norms scores compared to syphilis testing. |

**Supplementary Table 6 HIV positivity among people tested for HIV who attended an STI service**

| Author | Article | No of people tested for HIV | No of HIV positives | HIV Positivity (%) | Recruitment Site | Population |
|---|---|---|---|---|---|---|
| Assi A, 2019[3] | Prevalence of HIV and other sexually transmitted infections and their association with sexual practices and substance use among 2238 MSM in Lebanon | 2126 | 119 | 5.59 | STI Clinic | MSM |
| Banerjee, 2020[9] | A service evaluation comparing home-based testing to clinic-based testing for HIV, syphilis and hepatitis B in Birmingham and Solihull | 16229 | 42 | 0.25 | STI Clinic | Mixed |
| Bremer, 2016[17] | STI tests and proportion of positive tests in female sex workers attending local public health departments in Germany in 2010/11 | 3882 | 8 | 0.20 | STI Clinic | Sex workers |
| Chow, 2018[23] | Evaluation of the Implementation of a New Nurse-Led Express "Test-And-Go" Human Immunodeficiency Virus/Sexually Transmitted Infection Testing Service for Men Who Have Sex With Men at a Sexual Health Center in Melbourne, Australia | 3400 | 7 | 0.20 | STI Clinic | MSM |
| Kharsany, 2010[42] | Uptake of provider-initiated HIV testing and counseling among women attending an urban sexually transmitted disease clinic in South Africa – missed opportunities for early diagnosis of HIV infection | 2439 | 1378 | 56.49 | STI Clinic | Women attending STI clinics |

| | | | | | | |
|---|---|---|---|---|---|---|
| Katz, 2016[43] | Integrating HIV Testing as an Outcome of STD Partner Services for Men Who Have Sex with Men | 4441 | 104 | 2.34 | STI Clinic | MSM |
| Kilmarx, P. H, 2018[44] | HIV infection in patients with sexually transmitted infections in Zimbabwe - Results from the Zimbabwe STI etiology study | 489 | 201 | 41.10 | STI Clinic | Mixed |
| Leon, N, 2010[49] | The impact of provider-initiated (opt-out) HIV testing and counseling of patients with sexually transmitted infection in Cape Town, South Africa: a controlled trial | 1752 | 326 | 18.60 | STI Clinic | Mixed |
| Leon, N, 2010[49] | The impact of provider-initiated (opt-out) HIV testing and counseling of patients with sexually transmitted infection in Cape Town, South Africa: a controlled trial | 2821 | 605 | 21.44 | STI Clinic | Mixed |
| Llata, 2018[51] | New Human Immunodeficiency Virus Diagnoses Among Men Who Have Sex With Men Attending Sexually Transmitted Disease Clinics, STD Surveillance Network, January 2010 to June 2013 | 38915 | 640 | 1.64 | STI Clinic | MSM |
| Selvey, 2018[77] | Incidence and predictors of HIV, chlamydia and gonorrhoea among MSM attending a peer based clinic | 2753 | 46 | 1.67 | STI Clinic | MSM |
| Zhao, P.[95] | Uptake of provider-initiated HIV and syphilis testing among heterosexual STD clinic patients in Guangdong, China: results from a cross-sectional study | 1177 | 26 | 2.20 | STI Clinic | Mixed |

**Supplementary Table 7 HIV positivity among people who were tested for STIs**

| Author | Article | No of people tested for HIV | No of HIV positives | HIV Positivity (%) | Recruitment Site | Population |
|---|---|---|---|---|---|---|
| Balira, 2015[8] | The need for further integration of services to prevent mother-to-child transmission of HIV and syphilis in Mwanza City, Tanzania | 903 | 0 | 0 | Hospital<br>Reproductive and child health clinics | Health workers<br>Pregnant women |
| Carcamo, 2012[20] | Prevalences of sexually transmitted infections in young adults and female sex workers in Peru: a national population-based survey | 5681 | 22 | 0.38 | Outpatient | MSM |
| Carcamo, 2012[20] | Prevalences of sexually transmitted infections in young adults and female sex workers in Peru: a national population-based survey | 5799 | 3 | 0.05 | Outpatient | MSM |
| Kapadia, S. N, 2018[97] | Missed Opportunities for HIV Testing of Patients Tested for Sexually Transmitted Infections at a Large Urban Health Care System From 2010 to 2015 | 11515 | 31 | 0.26 | Hospital<br>Outpatient<br>Emergency Department | Male |
| Kapadia, S. N, 2018[97] | Missed Opportunities for HIV Testing of Patients Tested for Sexually Transmitted Infections at a Large Urban Health Care System From 2010 to 2015 | 56138 | 6 | 0.01 | Hospital<br>Outpatient | Female |

| | | | | | | |
|---|---|---|---|---|---|---|
| | | | | | Emergency Department | |
| Wood, 2014[92] | Outreach sexual infection screening and postal tests in men who have sex with men: are they comparable to clinic screening? | 30 | 1 | 3.33 | Other - Suana | MSM |
| Wood, 2014[92] | Outreach sexual infection screening and postal tests in men who have sex with men: are they comparable to clinic screening? | 30 | 0 | 0 | Other – Postal kits were sent | MSM |
| Wood, 2014[92] | Outreach sexual infection screening and postal tests in men who have sex with men: are they comparable to clinic screening? | 30 | 1 | 3.33 | STI Clinic | MSM |

**Supplementary Table 8 HIV positivity among people who were tested for STIs**

| Author | Article | No of people tested for HIV | No of HIV positives | HIV Positivity (%) | Recruitment Site | Population |
|---|---|---|---|---|---|---|
| Golden, 2015[28] | Sexually Transmitted Disease Partner Services Increase HIV Testing Among Men Who Have Sex with Men | 4631 | 165 | 3.56 | Other- partner service interviewed by public health staff | MSM |
| Cayuelas, 2019[21] | Indicator condition-guided HIV testing with an electronic prompt in primary healthcare: a before and after evaluation of an intervention | 34 | 2 | 5.88 | Community-based facility/GP | Mixed |
| Cayuelas, 2019[21] | Indicator condition-guided HIV testing with an electronic prompt in primary healthcare: a before and after evaluation of an intervention | 87 | 4 | 4.59 | Community-based facility/GP | Mixed |
| Cayuelas, 2019[21] | Indicator condition-guided HIV testing with an electronic prompt in primary healthcare: a before and after evaluation of an intervention | 73 | 3 | 4.10 | Community-based facility/GP | Mixed |
| Li, 2016[98] | HIV detection and prevalence among sexually transmitted diseases clinic patients in seven provinces (Autonomous Region) | 2668 | 63 | 2.36 | STI Clinic | Mixed |
| Petsis, 2020[69] | HIV Testing Among Adolescents With Acute Sexually Transmitted Infections | 1001 | 1 | 1 | Community based facility/GP | Youth |
| Williford, 2021[91] | HIV Screening Among Gonorrhea-Diagnosed Individuals; Baltimore, Maryland; April 2015 to April 2019 | 961 | 15 | 1.56 | STI Clinic | Mixed |

| | | | | | |
|---|---|---|---|---|---|
| | | | | | Emergency Department |

**Supplementary Table 9 HIV positivity among people with STI symptoms**

| Author | Article | No of people tested for HIV | No of HIV positives | HIV Positivity (%) | Recruitment Site | Population |
|---|---|---|---|---|---|---|
| Schechter, S. B, 2017[75] | Approach to Human Immunodeficiency Virus/Sexually Transmitted Infection Testing for Men at an Urban Urgent Care Center | 95 | 0 | 0 | Emergency Department | Males |
