## Additional File 3- Search Strategy for "Missed opportunities for HIV testing among those who accessed sexually transmitted infection (STI) services, tested for STIs and diagnosed with STIs: a systematic review and meta-analysis"

### SEARCH METHODOLOGY

For this systematic review, we searched five databases on May 5, 2021. Key terms for the search strategy included a combination of the following terms: “HIV infections/diagnosis [MeSH], “sexually transmitted diseases/diagnosis [MeSH]”. “HIV test”, “screening”, “mass screening [MeSH], and “missed opportunities”. The search limits were from 2000-current, humans, and English language. The search strategy was refined with the project team until the results retrieved reflected the scope of the project. The final Medline search was amended to run across the other databases. The databases searched were:

1. OvidSP MEDLINE and Epub Ahead of Print, In-Process & Other Non-Indexed Citations, Daily and Versions, 1946 to May 5, 2021
2. OvidSP Embase Classic + Embase, 1947 to May 5, 2021
3. Ovid Global Health, 1973 to 2021 week 21
4. EBSCO CINAHL Plus,
5. Web of Science Core Collection
    a. Science citation index expanded (1900- present)
    b. Social sciences citation index (1900 to present)
    c. Arts & humanities citation index (1975 to present)
    d. Conference proceedings citation index- Science (1990 to present)
    e. Conference proceedings citation index - Social science & humanities (1990 to present)
    f. Book citation index - science (2005 to present)
    g. Book citation index - Social sciences & humanities (2005 to present)
    h. Current chemical reactions (1985 to present)
    i. Index Chemicus (1993 to present).

### SEARCH RESULTS

| Database name | EndNote import order | Number of references before deduplication | Number of references after deduplication (removed) |
| --- | --- | --- | --- |
| Medline | 1 | 3004 | 2999 |
| Embase | 2 | 4299 | 4206 |
| Global Health | 3 | 3126 | 3126 |
| CINAHL | 4 | 1,444 | 1,412 |
| Web of Science | 6 | 2,583 | 2,580 |
| **Total** | | 14,456 | 14323<br>After deduplication against all in Endnote= 9726 |

### Search Strategies

#### 1.1 OvidSP Medline

| | |
| --- | --- |
| Database name | Medline |
| Database platform | OvidSP |

| | |
|---|---|
| Dates of database coverage | 1946 to May 5 2021 |
| Date searched | 07/05/2021 |
| Searched by | KS |
| Number of hits | 3004 |

| # | Searches | Results |
|---|---|---|
| 1 | sexually transmitted diseases/ or sexually transmitted diseases, bacterial/ or chancroid/ or chlamydia infections/ or lymphogranuloma venereum/ or gonorrhea/ or granuloma inguinale/ or syphilis/ or sexually transmitted diseases, viral/ or condylomata acuminata/ or herpes genitalis/ | 82501 |
| 2 | ((sexual* transmi* or venereal) adj (disease* or infection*)).mp. | 48311 |
| 3 | (Chancroid* or Ducrey disease or H?emophilus ducrey* infection* or ulcus molle or Donovanosis or Condyloma acuminat* or condyla acuminat* or verruca accuminata or vulvar condyloma or Trichomoniasis or gonorrh?ea* or granuloma venereum or lymphogranuloma venereum or herpes genitalis or herpes progenitalis or herpes simplex genitalis or progenitalis,herpes or syphilis or syphilitic chancre or gonococcal urethritis or gonorrh?ic urethritis or urethritis gonorrhoica).mp. | 68875 |
| 4 | ((genital* or anogenital* or venereal or anal or perianal or penile or vulvar or perigenital* or urogenital) adj (wart? or herpes)).mp. | 6532 |
| 5 | (sexual health adj (clinic* or unit* or service* or department*)).mp. | 1650 |

| | | |
|---|---|---|
| 6 | 1 or 2 or 3 or 4 or 5 | 121701 |
| 7 | HIV/ or HIV-2/ or HIV-1/ | 101473 |
| 8 | exp HIV Infections/ | 290992 |
| 9 | (human immun* deficienc* or human immunodeficienc* or acquired immun* deficienc* or acquired immunodeficienc* or HIV).mp. | 420910 |
| 10 | (HIV adj2 AIDS).mp. | 37461 |
| 11 | 7 or 8 or 9 or 10 | 425722 |
| 12 | 6 and 11 | 25013 |
| 13 | early diagnosis/ | 27831 |
| 14 | Mass Screening/ | 107183 |
| 15 | Delayed Diagnosis/ | 6935 |
| 16 | 13 or 14 or 15 | 139527 |
| 17 | 12 and 16 | 1373 |
| 18 | hiv testing/ or aids serodiagnosis/ | 6882 |
| 19 | 6 and 18 | 489 |
| 20 | exp HIV Infections/di [Diagnosis] | 31540 |
| 21 | 6 and 20 | 2382 |
| 22 | ((screen* or test* or early diagnos* or early detect*) adj2 (HIV or human immun* deficienc* or human immunodeficienc* or acquired immun* deficienc* or acquired immunodeficienc*)).mp. | 25995 |

| | | |
|---|---|---|
| 23 | ((missed adj2 opportunit*) and (HIV or human immun* deficienc* or human immunodeficienc* or acquired immun* deficienc* or acquired immunodeficienc*)).mp. | 653 |
| 24 | 22 or 23 | 26333 |
| 25 | 6 and 24 | 3973 |
| 26 | 17 or 19 or 21 or 25 | 5865 |
| 27 | limit 26 to yr="2010 -Current" | 3277 |
| 28 | exp animals/ not humans.sh. | 4821824 |
| 29 | 27 not 28 | 3276 |
| 30 | limit 29 to (case reports or comment or editorial or letter or news or newspaper article or "systematic review") | 272 |
| 31 | 29 not 30 | 3004 |

#### 1.2 Embase Classic + Embase

| | |
|---|---|
| Database name | Embase Classic + Embase |
| Database platform | OvidSP |
| Dates of database coverage | 1946 to May 5 2021 |
| Date searched | 07/05/2021 |
| Searched by | KS |
| Number of hits | 4299 |

| # | Searches | Results |
|---|---|---|
| 1 | sexually transmitted diseases/ or sexually transmitted diseases, bacterial/ or chancroid/ or chlamydia infections/ or lymphogranuloma venereum/ or gonorrhea/ or granuloma inguinale/ or syphilis/ or sexually transmitted diseases, viral/ or condylomata acuminata/ or herpes genitalis/ | 109090 |
| 2 | ((sexual* transmi* or venereal) adj (disease* or infection*)).mp. | 69677 |
| 3 | (Chancroid* or Ducrey disease or H?emophilus ducrey* infection* or ulcus molle or Donovanosis or Condyloma acuminat* or condyla acuminat* or verruca accuminata or vulvar condyloma or Trichomoniasis or gonorrh?ea* or granuloma venereum or lymphogranuloma venereum or herpes genitalis or herpes progenitalis or herpes simplex genitalis or progenitalis,herpes or syphilis or syphilitic chancre or gonococcal urethritis or gonorrh?ic urethritis or urethritis gonorrhoica).mp. | 102476 |

| | | |
|---|---|---|
| 4 | ((genital* or anogenital* or venereal or anal or perianal or penile or vulvar or perigenital* or urogenital) adj (wart? or herpes)).mp. | 11983 |
| 5 | (sexual health adj (clinic* or unit* or service* or department*)).mp. | 3200 |
| 6 | 1 or 2 or 3 or 4 or 5 | 163512 |
| 7 | HIV/ or HIV-2/ or HIV-1/ | 197828 |
| 8 | exp HIV Infections/ | 394901 |
| 9 | (human immun* deficienc* or human immunodeficienc* or acquired immun* deficienc* or acquired immunodeficienc* or HIV).mp. | 584823 |
| 10 | (HIV adj2 AIDS).mp. | 47272 |
| 11 | 7 or 8 or 9 or 10 | 586432 |
| 12 | 6 and 11 | 40723 |
| 13 | early diagnosis/ | 122506 |
| 14 | Mass Screening/ | 62604 |
| 15 | Delayed Diagnosis/ | 14266 |
| 16 | 13 or 14 or 15 | 195690 |
| 17 | 12 and 16 | 1127 |
| 18 | hiv testing/ or aids serodiagnosis/ | 59369 |
| 19 | 6 and 18 | 4828 |
| 20 | exp HIV Infections/di [Diagnosis] | 34045 |

| | | |
|---|---|---|
| 21 | 6 and 20 | 3166 |
| 22 | ((screen* or test* or early diagnos* or early detect*) adj2 (HIV or human immun* deficienc* or human immunodeficienc* or acquired immun* deficienc* or acquired immunodeficienc*)).mp. | 38144 |
| 23 | ((missed adj2 opportunit*) and (HIV or human immun* deficienc* or human immunodeficienc* or acquired immun* deficienc* or acquired immunodeficienc*)).mp. | 1011 |
| 24 | 22 or 23 | 38630 |
| 25 | 6 and 24 | 6652 |
| 26 | 17 or 19 or 21 or 25 | 11382 |
| 27 | limit 26 to yr="2010 -Current" | 6083 |
| 28 | (exp animal/ or nonhuman/) not exp human/ | 7449210 |
| 29 | 27 not 28 | 6064 |
| 30 | limit 29 to (editorial or letter or note or "review") | 674 |
| 31 | 29 not 30 | 5390 |
| 32 | (case report* or editorial* or letter* or news* or review*).mp. | 9038279 |
| 33 | 31 not 32 | 4299 |

#### 1.3 GLOBAL HEALTH

| | |
|---|---|
| Database name | Global Health |
| Database platform | OvidSP |
| Dates of database coverage | 2nd Quarter 2021 |
| Date searched | 07/05/2021 |
| Searched by | KS |
| Number of hits | 3126 |

| # | Searches | Results |
|---|---|---|
| 1 | sexually transmitted diseases/ or sexually transmitted diseases, bacterial/ or chancroid/ or chlamydia infections/ or lymphogranuloma venereum/ or gonorrhea/ or granuloma inguinale/ or syphilis/ or sexually transmitted diseases, viral/ or condylomata acuminata/ or herpes genitalis/ | 36905 |
| 2 | ((sexual* transmi* or venereal) adj (disease* or infection*)).mp. | 37666 |
| 3 | (Chancroid* or Ducrey disease or H?emophilus ducrey* infection* or ulcus molle or Donovanosis or Condyloma acuminat* or condyla acuminat* or verruca accuminata or vulvar condyloma or Trichomoniasis or gonorrh?ea* or granuloma venereum or lymphogranuloma venereum or herpes genitalis or herpes progenitalis or herpes simplex genitalis or progenitalis,herpes or syphilis or syphilitic chancre or gonococcal urethritis or gonorrh?ic urethritis or urethritis gonorrhoica).mp. | 21132 |

| | | |
|---|---|---|
| 4 | ((genital* or anogenital* or venereal or anal or perianal or penile or vulvar or perigenital* or urogenital) adj (wart? or herpes)).mp. | 3092 |
| 5 | (sexual health adj (clinic* or unit* or service* or department*)).mp. | 1083 |
| 6 | 1 or 2 or 3 or 4 or 5 | 48926 |
| 7 | HIV/ or HIV-2/ or HIV-1/ | 162642 |
| 8 | exp HIV Infections/ | 152488 |
| 9 | (human immun* deficienc* or human immunodeficienc* or acquired immun* deficienc* or acquired immunodeficienc* or HIV).mp. | 200531 |
| 10 | (HIV adj2 AIDS).mp. | 22974 |
| 11 | 7 or 8 or 9 or 10 | 200531 |
| 12 | 6 and 11 | 24488 |
| 13 | early diagnosis/ | 2153 |
| 14 | (delayed diagnos* or diagnostic dela*).mp. | 1858 |
| 15 | mass screeni*.mp. | 912 |
| 16 | 13 or 14 or 15 | 4892 |
| 17 | 12 and 16 | 72 |
| 18 | (hiv testin* or hiv screenin* or aids serodiagnos*).mp. | 8719 |
| 19 | 6 and 18 | 2328 |

| | | |
|---|---|---|
| 20 | ((screen* or test* or early diagnos* or early detect*) adj2 (HIV or human immun* deficienc* or human immunodeficienc* or acquired immun* deficienc* or acquired immunodeficienc*)).mp. | 20108 |
| 21 | ((missed adj2 opportunit*) and (HIV or human immun* deficienc* or human immunodeficienc* or acquired immun* deficienc* or acquired immunodeficienc*)).mp. | 438 |
| 22 | 20 or 21 | 20305 |
| 23 | 6 and 22 | 5164 |
| 24 | 17 or 19 or 23 | 5196 |
| 25 | limit 24 to (bulletin or correspondence or editorial or thesis or miscellaneous) | 134 |
| 26 | 24 not 25 | 5062 |
| 27 | (case report* or editorial* or letter* or news*).mp. | 110237 |
| 28 | 26 not 27 | 4973 |
| 29 | (mice* or rat or rats or dog or dogs or sheep* or pig* or rodents).mp. | 499700 |
| 30 | 28 not 29 | 4969 |
| 31 | limit 30 to (english language and yr="2010 -Current") | 3126 |

### 1.4 CINAHL PLUS

| | |
|---|---|
| Database name | CINAHL PLUS |
| Database platform | EBSCO |
| Dates of database coverage | |
| Date searched | 07/05/2021 |
| Searched by | KS |
| Number of hits | 1,444 |

| # | Searches | Results |
|---|---|---|
| 1 | (MH "Sexually Transmitted Diseases") OR (MH "Sexually Transmitted Diseases, Viral") OR (MH "Sexually Transmitted Diseases, Protozoal") OR (MH "Sexually Transmitted Diseases, Fungal") OR (MH "Sexually Transmitted Diseases, Bacterial") | 14,252 |
| 2 | (("sexual* transmi*" or venereal) N2 (disease* or infection*)) | 20,534 |
| 3 | (Chancroid* or "Ducrey disease" or "H#emophilus ducrey* infection*" or "ulcus molle" or Donovanosis or "Condyloma acuminat*" or "condyla acuminat*" or "verruca accuminata" or "vulvar condyloma" or Trichomoniasis or gonorrh#ea* or "granuloma venereum" or "lymphogranuloma venereum" or "herpes genitalis" or "herpes progenitalis" or "herpes simplex genitalis" or "progenitalis herpes" or syphilis or "syphilitic chancre" or "gonococcal urethritis" or "gonorrh#ic urethritis" or "urethritis gonorrhoica") | 11,547 |
| 4 | ((genital* or anogenital* or venereal or anal or perianal or penile or vulvar or perigenital* or urogenital) N2 (wart# or herpes)) | 3,243 |

| | | |
|---|---|---|
| 5 | (("sexual health") N2 (clinic* or unit* or service* or department*)) | 1,460 |
| 6 | S1 OR S2 OR S3 OR S4 OR S5 OR S6 | 30,844 |
| 7 | (MH "Human Immunodeficiency Virus") | 5,314 |
| 8 | (MH "HIV-1") | 5,364 |
| 9 | (MH "HIV Infections") | 70,836 |
| 10 | (("human immun* deficienc*") or ("acquired immun* deficienc*") or (HIV)) | 119,229 |
| 11 | S7 OR S8 OR S9 OR S10 | 119,498 |
| 12 | S6 AND S11 | 10,435 |
| 13 | (MH "Early Diagnosis+") | 20,888 |
| 14 | (MH "Diagnosis, Delayed") | 4,550 |
| 15 | (MH "Mass Screening") | 1,130 |
| 16 | (("early diagnos*" or "early intervention") or ("delayed diagnosi*" or "diagnostic dela*") or ("mass screenin*")) | 46,980 |
| 17 | S13 OR S14 OR S15 OR S16 | 58,521 |
| 18 | S12 AND S17 | 155 |
| 19 | (("hiv testin*" or "hiv screenin*") or ("aids serodiagnos*")) | 8,988 |
| 20 | ("hiv infection diagnos*") | 64 |
| 21 | S19 OR S20 | 9,033 |
| 22 | S6 AND S21 | 1,025 |

| | | |
|---|---|---|
| 23 | ((screen* or test* or "early diagnos*" or "early detect*") N2 (HIV or "human immun* deficienc*" or "human immunodeficienc*" or "acquired immun* deficienc*" or "acquired immunodeficienc*")) | 11,690 |
| 24 | ((missed N2 opportunit*) and (HIV or "human immun* deficienc*" or "human immunodeficienc*" or "acquired immun* deficienc*" or "acquired immunodeficienc*")) | 315 |
| 25 | S23 OR S24 | 11,849 |
| 26 | S6 AND S25 | 1,834 |
| 27 | S18 OR S22 OR S26 | 2,047 |
| 28 | (animal* not human*) | 178,571 |
| 29 | S27 not S28<br>**Limiters** - Publication Year: 2010-2021; English Language | 1,444 |

#### 1.5 Web of Science

| | |
|---|---|
| Database name | Web of Science Core Collection |
| Database platform | Web of Science |
| Dates of database coverage | Science citation index expanded (1900- present)<br>Social sciences citation index (1900 to present)<br>Arts & humanities citation index (1975 to present)<br>Conference proceedings citation index- Science (1990 to present)<br>Conference proceedings citation index - Social science & humanities (1990 to present)<br>Book citation index - science (2005 to present)<br>Book citation index - Social sciences & humanities (2005 to present)<br>Current chemical reactions (1985 to present)<br>Index Chemicus (1993 to present) |
| Date searched | 07/05/2021 |
| Searched by | KS |
| Number of hits | 2,583 |

| # | Searches | Results |
|---|---|---|
| 1 | TS=(("sexual* transmi*" or venereal) NEAR/0 (disease* or infection*) ) | 37,407 |

| | | |
|---|---|---|
| 2 | TS=(Chancroid* or "Ducrey disease*" or "H$emophilus ducrey* infection*" or "ulcus molle" or Donovanosis or "Condyloma acuminat*" or "condyla acuminat*" or "verruca accuminata" or "vulvar condyloma" or Trichomoniasis or gonorrh$ea* or "granuloma venereum" or "lymphogranuloma venereum" or "herpes genitalis" or "herpes progenitalis" or "herpes simplex genitalis" or "progenitalis herpes" or syphilis or "syphilitic chancre" or "gonococcal urethritis" or "gonorrh$ic urethritis" or "urethritis gonorrhoica") | 49,211 |
| 3 | TS=((genital* or anogenital* or venereal or anal or perianal or penile or vulvar or perigenital* or urogenital) NEAR/0 (wart$ or herpes) ) | 8,128 |
| 4 | TS=(("sexual health") NEAR/0 (clinic* or unit* or service* or department*) ) | 2,067 |
| 5 | #4 OR #3 OR #2 OR #1 | 84,461 |
| 6 | TS=(HIV or "HIV-2" or "HIV-1") | 395,643 |
| 7 | TS= ("HIV Infection*") | 79,798 |
| 8 | TS=("human immun* deficienc*" or "human immunodeficienc*" or "acquired immun* deficienc*" or "acquired immunodeficienc*" or "HIV") | 435,910 |
| 9 | TS=((HIV) NEAR/2 (AIDS) ) | 48,956 |
| 10 | #9 OR #8 OR #7 OR #6 | 435,910 |
| 11 | #5 AND #10 | 23,221 |
| 12 | TS=("early diagnos*" or "mass screenin*" or "delayed diagnos*") | 94,245 |
| 13 | #11 AND #12 | 213 |
| 14 | TS=(("hiv testin*" or "hiv screening*") or ("aids serodiagnos*") ) | 12,871 |
| 15 | TS=("HIV Infectio* diagnos*") | 245 |

| | | |
|---|---|---|
| 16 | #15 OR #14 | 13,060 |
| 17 | #16 AND #5 | 1,890 |
| 18 | TS=((screen* or test* or "early diagnos*" or "early detect*") NEAR/2 (HIV or "human immun* deficienc*" or "human immunodeficienc*" or "acquired immun* deficienc*" or "acquired immunodeficienc*") ) | 28,048 |
| 19 | TS=((missed NEAR/2 opportunit*) and (HIV or "human immun* deficienc*" or "human immunodeficienc*" or "acquired immun* deficienc*" or "acquired immunodeficienc*") ) | 872 |
| 20 | #19 OR #18 | 28,488 |
| 21 | #20 AND #5 | 4,205 |
| 22 | #21 OR #17 OR #13 | 4,361 |
| 23 | #21 OR #17 OR #13<br>**Refined by:** [excluding] **DOCUMENT TYPES:** ( LETTER OR BOOK CHAPTER OR REVIEW OR EDITORIAL MATERIAL OR NOTE )<br>*Indexes=SCI-EXPANDED, SSCI, A&HCI, CPCI-S, CPCI-SSH, BKCI-S, BKCI-SSH, ESCI, CCR-EXPANDED, IC*<br>*Timespan=All years* | 4,077 |
| 24 | #21 OR #17 OR #13<br>**Refined by: PUBLICATION YEARS:** ( 2021 OR 2012 OR 2020 OR 2011 OR 2019 OR 2010 OR 2018 OR 2017 OR 2016 OR 2015 OR 2014 )<br>*Indexes=SCI-EXPANDED, SSCI, A&HCI, CPCI-S, CPCI-SSH, BKCI-S, BKCI-SSH, ESCI, CCR-EXPANDED, IC*<br>*Timespan=All years* | 2,617 |
| 25 | #21 OR #17 OR #13 | 2,583 |

| | |
|---|---|
| | **Refined by: PUBLICATION YEARS:** ( 2021 OR 2012 OR 2020 OR 2011 OR 2019 OR 2010 OR 2018 OR 2017 OR 2016 OR 2015 OR 2014 ) AND [excluding] **WEB OF SCIENCE CATEGORIES:** ( ENGINEERING BIOMEDICAL OR GEOGRAPHY OR INFORMATION SCIENCE LIBRARY SCIENCE OR MATHEMATICAL COMPUTATIONAL BIOLOGY OR BIOTECHNOLOGY APPLIED MICROBIOLOGY OR BIOCHEMICAL RESEARCH METHODS OR CELL TISSUE ENGINEERING OR CHEMISTRY APPLIED OR CHEMISTRY MEDICINAL OR DEMOGRAPHY OR FOOD SCIENCE TECHNOLOGY OR HISTORY PHILOSOPHY OF SCIENCE OR CRIMINOLOGY PENOLOGY OR INTEGRATIVE COMPLEMENTARY MEDICINE OR NUTRITION DIETETICS OR ZOOLOGY OR VETERINARY SCIENCES )<br>*Indexes=SCI-EXPANDED, SSCI, A&HCI, CPCI-S, CPCI-SSH, BKCI-S, BKCI-SSH, ESCI, CCR-EXPANDED, IC*<br>*Timespan=All years* |
