## Additional File 4- Quality Checklists for "Missed opportunities for HIV testing among those who accessed sexually transmitted infection (STI) services, tested for STIs and diagnosed with STIs: a systematic review and meta-analysis"

**CHEERS Checklist- Modelling Studies**

| | Beck, 2016[1] | Owusu-Edusei, 2014[2] | Prabhu, 2011[3] |
|---|---|---|---|
| Title | 1 | 1 | 1 |
| Abstract | 1 | 1 | 1 |
| Background and objectives | 0 | 1 | 1 |
| Target population and subgroups | 1 | 1 | 1 |
| Setting and location | 1 | 1 | 1 |
| Study perspective | 0 | 1 | 1 |
| Comparators | 1 | 1 | 1 |
| Time horizon | 1 | 0 | 0 |
| Discount rate | 0 | 1 | 1 |

| | | | |
|---|---|---|---|
| Choice of health outcomes | 1 | 1 | 1 |
| Measurement of effectiveness | 0 | 1 | 1 |
| Measurement and valuation of preference-based outcomes | 0 | 1 | 1 |
| Estimating resources and costs | 0 | 1 | 1 |
| Currency, price date, and conversion | 0 | 1 | 1 |
| Choice of model | 1 | 1 | 1 |
| Assumptions | 1 | 1 | 1 |
| Analytical methods | 0 | 1 | 1 |
| Study parameters | 0 | 1 | 1 |
| Incremental costs and outcomes | 1 | 1 | 1 |
| Characterising uncertainty | 0 | 1 | 1 |

| | | | |
|---|---|---|---|
| Characterising heterogeneity | 0 | 1 | 1 |
| Study findings, limitations,generalisability, and current knowledge | 0 | 1 | 1 |
| Source of funding | 0 | 1 | 1 |
| Conflicts of interest | 0 | 1 | 1 |
| **Total score (Out of 24)** | 9 | 20 | 21 |

ROB 2.0 – RCT

| Authors | Bauermeister, 2015[4] | Brown, 2010[5] | Sharma, 2019[6] |
|---|---|---|---|
| Domain 1: Risk of bias arising from the randomization process | | | |
| 1.1 Was the allocation sequence random? | Y | Y | NI |
| 1.2 Was the allocation sequence concealed until participants were enrolled and assigned to interventions? | Y | Y | Y |
| 1.3 Did baseline differences between intervention groups suggest a problem with the randomization process? | N | N | N |
| Risk-of-bias judgement | Low risk | Low risk | Some concerns |

| Domain 2: Risk of bias due to deviations from the intended interventions (effect of assignment to intervention) | | | |
|---|---|---|---|
| 2.1 Were participants aware of their assigned intervention during the trial? | N | N | N |
| 2.2 Were carers and people delivering the interventions aware of participants' assigned intervention during the trial? | N | N | N |
| 2.3 If Y/PY/NI to 2.1 or 2.2: Were there deviations from the intended intervention that arose because of the trial context? | NA | NA | NA |
| 2.4 If Y/PY to 2.3: Were these deviations likely to have affected the outcome? | NA | NA | NA |

| | | | |
|---|---|---|---|
| 2.5 If Y/PY/NI to 2.4: Were these deviations from intended intervention balanced between groups? | NA | NA | NA |
| 2.6 Was an appropriate analysis used to estimate the effect of assignment to intervention? | Y | Y | Y |
| 2.7 If N/PN/NI to 2.6: Was there potential for a substantial impact (on the result) of the failure to analyse participants in the group to which they were randomized? | NA | NA | NA |
| Risk-of-bias judgement | Low risk | Low risk | Low risk |
| Domain 3: Risk of bias due to missing outcome data | | | |
| | Y | Y | Y |

| | | | |
|---|---|---|---|
| 3.1 Were data for this outcome available for all, or nearly all, participants randomized? | | | |
| 3.2 If N/PN/NI to 3.1: Is there evidence that the result was not biased by missing outcome data? | NA | NA | NA |
| 3.3 If N/PN to 3.2: Could missingness in the outcome depend on its true value? | NA | NA | NA |
| 3.4 If Y/PY/NI to 3.3: Is it likely that missingness in the outcome depended on its true value? | NA | NA | NA |
| Risk-of-bias judgement | Low risk | Low risk | Low risk |

| Domain 4: Risk of bias in measurement of the outcome | | | |
|---|---|---|---|
| 4.1 Was the method of measuring the outcome inappropriate? | N | N | N |
| 4.2 Could measurement or ascertainment of the outcome have differed between intervention groups? | N | N | N |
| 4.3 If N/PN/NI to 4.1 and 4.2: Were outcome assessors aware of the intervention received by study participants? | N | N | N |
| 4.4 If Y/PY/NI to 4.3: Could assessment of the outcome have been influenced by knowledge of intervention received? | NA | NA | NA |

| | | | |
|---|---|---|---|
| 4.5 If Y/PY/NI to 4.4: Is it likely that assessment of the outcome was influenced by knowledge of intervention received? | NA | NA | NA |
| Risk-of-bias judgement | Low risk | Low risk | Low risk |
| Domain 5: Risk of bias in selection of the reported result | | | |
| 5.1 Were the data that produced this result analysed in accordance with a pre-specified analysis plan that was finalized before unblinded outcome data were available for analysis? | Y | Y | Y |
| 5.2 Is the numerical result being assessed likely to have been selected, on the basis of the results, from multiple eligible outcome measurements (e.g., scales, | N | N | N |

| | | | |
|---|---|---|---|
| definitions, time points) within the outcome domain? | | | |
| 5.3 Is the numerical result being assessed likely to have been selected, on the basis of the results, from multiple eligible analyses of the data? | N | N | N |
| Risk-of-bias judgement | Low risk | Low risk | Low risk |
| Overall risk of bias (Low / High / Some concerns) | **Low risk** | **Low risk** | **Some concerns** |

**Newcastle-Ottawa Scale - Cohort Studies**

| **Authors** | Representativeness of the exposed cohort<br><br>a) truly representative of the average _______________ (describe) in the community<br><br>b) somewhat representative of the average ______________ in the community<br><br>c) selected group of users eg nurses, volunteers<br><br>d) no description of the derivation of the cohort | Selection of the non exposed cohort<br><br>a) drawn from the same community as the exposed cohort<br><br>b) drawn from a different source<br><br>c) no description of the derivation of the non exposed cohort | Ascertainment of exposure<br><br>a) secure record (eg surgical records)<br><br>b) structured interview<br><br>c) written self report<br><br>d) no description | Demonstration that outcome of interest was not present at start of study<br><br>a) yes<br><br>b) no | Comparability of cohorts on the basis of the design or analysis<br><br>a) study controls for _____________ (select the most important factor)<br><br>b) study controls for any additional factor | Assessment of outcome<br><br>a) independent blind assessment<br><br>b) record linkage<br><br>c) self report<br><br>d) no description | Was follow-up long enough for outcomes to occur<br><br>a) yes (select an adequate follow up period for outcome of interest)<br><br>b) no | Adequacy of follow up of cohorts<br><br>a) complete follow up - all subjects accounted for<br><br>b) subjects lost to follow up unlikely to introduce bias - small number lost - > ____ % (select an adequate %) follow up, or description provided of those lost)<br><br>c) follow up rate < ____% (select an adequate %) and no | Score |
|---|---|---|---|---|---|---|---|---|---|

| | | | | | | | | description of those lost<br><br>d) no statement | |
|---|---|---|---|---|---|---|---|---|---|
| Cayuelas, 2019[7] | 1 | 1 | 1 | 1 | 2 | 1 | 0 | 1 | 8 |
| Fernandez, 2016[8] | 1 | 0 | 0 | 1 | 2 | 0 | 0 | 1 | 5 |
| Golden, 2015[9] | 1 | 0 | 1 | 1 | 1 | 1 | 0 | 0 | 5 |
| Goulet, 2014[10] | 1 | 0 | 1 | 1 | 2 | 1 | 1 | 1 | 8 |
| Jichlinski, 2018[11] | 1 | 0 | 1 | 1 | 2 | 1 | 1 | 1 | 8 |
| Joore, 2016[12] | 1 | 0 | 1 | 1 | 2 | 0 | 1 | 1 | 6 |
| Kapadia, 2018[13] | 1 | 0 | 1 | 1 | 2 | 1 | 0 | 1 | 7 |
| Katz, 2016[14] | 1 | 0 | 1 | 1 | 2 | 1 | 0 | 1 | 7 |
| Klein, 2011[15] | 1 | 0 | 1 | 1 | 2 | 1 | 0 | 1 | 7 |

| | | | | | | | | | |
|---|---|---|---|---|---|---|---|---|---|
| Lanier, 2014[16] | 1 | 0 | 0 | 1 | 1 | 0 | 0 | 0 | 3 |
| Mullens, 2019[17] | 1 | 0 | 1 | 1 | 2 | 1 | 0 | 1 | 7 |
| Owusu-Edusei Jr, 2015[18] | 1 | 0 | 0 | 1 | 2 | 0 | 0 | 0 | 4 |
| Petlo, 2011[19] | 1 | 0 | 1 | 1 | 2 | 1 | 0 | 1 | 7 |
| Petsis, 2020[20] | 1 | 0 | 1 | 1 | 2 | 1 | 0 | 0 | 6 |
| Ruutel, 2018[21] | 1 | 0 | 1 | 1 | 2 | 1 | 0 | 1 | 7 |
| Schechter, 2017[22] | 1 | 0 | 1 | 1 | 2 | 1 | 1 | 1 | 8 |
| Selvey, 2018[23] | 1 | 0 | 1 | 1 | 2 | 1 | 1 | 1 | 8 |
| Snow, 2011[24] | 1 | 0 | 1 | 1 | 2 | 1 | 1 | 1 | 8 |
| Wang, 2008[25] | 1 | 1 | 1 | 1 | 2 | 1 | 1 | 1 | 9 |

| | | | | | | | | | |
|---|---|---|---|---|---|---|---|---|---|
| Ward, 2016[26] | 1 | 0 | 0 | 1 | 2 | 0 | 0 | 1 | 5 |
| Waxman, 2016[27] | 1 | 0 | 1 | 1 | 2 | 1 | 1 | 1 | 8 |
| Wood, 2014[28] | 1 | 0 | 1 | 1 | 1 | 1 | 0 | 0 | 5 |
| Youssef, 2018[29] | 1 | 1 | 1 | 1 | 2 | 1 | 1 | 1 | 9 |
| Yumori, 2021[30] | 1 | 0 | 1 | 1 | 2 | 1 | 0 | 1 | 7 |

**Newcastle-Ottawa Scale – Cross-sectional Studies**

| **Authors** | Representativeness of the sample:<br><br>a. Truly representative of the average in the target population. * (all subjects or random sampling)<br><br>b. Somewhat representative of the average in the target group. * (non-random sampling)<br><br>c. Selected group of users/convenience sample.<br><br>d. No description of the derivation of the included subjects. | Sample size :<br>a. Justified and satisfactory (including sample size calculation). *<br><br>b. Not justified.<br><br>c. No information provided | Non-respondents:<br><br>a. Proportion of target sample recruited attains pre-specified target or basic summary of non-respondent characteristics in sampling frame recorded. *<br><br>b. Unsatisfactory recruitment rate, no summary data on non-respondents.<br><br>c. No information provided | Ascertainment of the exposure (risk factor):<br><br>a. Vaccine records/vaccine registry/clinic registers/hospital records only. **<br><br>b. Parental or personal recall and vaccine/hospital records. *<br><br>c. Parental/personal recall only. | Comparability of subjects in different outcome groups on the basis of design or analysis. Confounding factors controlled.<br><br>a. Data/ results adjusted for relevant predictors/risk factors/confounders e.g. age, sex, time since vaccination, etc. **<br><br>b. Data/results not adjusted for all relevant confounders/risk factors/information not provided. | Assessment of outcome:<br><br>a. Independent blind assessment using objective validated laboratory methods. **<br><br>b. Unblinded assessment using objective validated laboratory methods. **<br><br>c. Used non-standard or non-validated laboratory methods with gold standard. *<br><br>d. No description/non-standard | Statistical test:<br><br>a. Statistical test used to analyse the data clearly described, appropriate and measures of association presented including confidence intervals and probability level (p value). *<br><br>b. Statistical test not appropriate, not described or incomplete. | Score |
|---|---|---|---|---|---|---|---|---|

| | | | | | | | | |
|---|---|---|---|---|---|---|---|---|
| | | | | | | laboratory methods used. | | |
| Adam, 2014[31] | 1 | 1 | 1 | 1 | 2 | 0 | 1 | 7 |
| Adekeye, 2016[32] | 1 | 1 | 0 | 2 | 2 | 0 | 1 | 7 |
| Assi, 2019[33] | 1 | 1 | 1 | 1 | 2 | 2 | 1 | 9 |
| Avoundjian, 2019[34] | 1 | 1 | 0 | 2 | 0 | 2 | 1 | 7 |
| Badman, 2016[35] | 1 | 1 | 0 | 2 | 2 | 2 | 1 | 9 |
| Baker, 2015[36] | 1 | 1 | 1 | 1 | 0 | 0 | 1 | 5 |
| Balira, 2015[37] | 1 | 1 | 1 | 1 | 0 | 1 | 0 | 5 |
| Banerjee, 2020[38] | 1 | 1 | 1 | 2 | 0 | 1 | 1 | 7 |
| Barber, 2011[39] | 1 | 1 | 1 | 1 | 0 | 0 | 1 | 5 |

| | | | | | | | | |
|---|---|---|---|---|---|---|---|---|
| Barnes, 2019[40] | 1 | 1 | 1 | 2 | 2 | 0 | 1 | 8 |
| Bauermeister, 2015[41] | 0 | 1 | 0 | 1 | 0 | 0 | 1 | 3 |
| Bradley, 2013[42] | 1 | 1 | 1 | 1 | 2 | 0 | 1 | 7 |
| Bremer, 2016[43] | 1 | 1 | 1 | 1 | 0 | 0 | 1 | 5 |
| Bristow, 2018[44] | 0 | 1 | 0 | 0 | 1 | 1 | 1 | 4 |
| Carcamo, 2012[45] | 1 | 1 | 1 | 2 | 2 | 2 | 1 | 10 |
| Chen J, 2011[46] | 1 | 1 | 0 | 2 | 2 | 0 | 1 | 7 |
| Chow E, 2018[47] | 1 | 1 | 1 | 2 | 2 | 2 | 1 | 10 |
| Cushman, 2019[48] | 1 | 1 | 1 | 1 | 0 | 0 | 1 | 5 |
| Gamagedara, 2014[49] | 1 | 1 | 1 | 2 | 2 | 2 | 1 | 10 |
| Gilbert, 2018[50] | 1 | 1 | 1 | 2 | 2 | 0 | 1 | 8 |

| | | | | | | | | |
|---|---|---|---|---|---|---|---|---|
| Goyal, 2013[51] | 1 | 1 | 1 | 2 | 2 | 2 | 1 | 10 |
| Heard, 2020[52] | 1 | 1 | 1 | 1 | 0 | 0 | 1 | 5 |
| Inghels, 2020[53] | 1 | 1 | 1 | 1 | 2 | 0 | 1 | 7 |
| Josten, 2018[54] | 1 | 0 | 0 | 1 | 0 | 0 | 0 | 2 |
| Kharsany, 2010[55] | 1 | 1 | 1 | 1 | 0 | 0 | 0 | 4 |
| Kilmarx, 2018[56] | 1 | 1 | 0 | 1 | 2 | 2 | 1 | 8 |
| Klein, 2014[57] | 1 | 1 | 1 | 2 | 2 | 2 | 1 | 10 |
| Li, J, 2016[58] | 1 | 1 | 0 | 1 | 0 | 2 | 1 | 6 |
| Llata, 2018[59] | 1 | 1 | 1 | 2 | 2 | 2 | 1 | 10 |

| | | | | | | | | |
|---|---|---|---|---|---|---|---|---|
| Lopez, 2019[60] | 1 | 1 | 0 | 2 | 0 | 2 | 1 | 7 |
| MacDonald, 2010[61] | 1 | 1 | 1 | 2 | 0 | 0 | 0 | 5 |
| Marsh, 2013[62] | 1 | 1 | 0 | 2 | 0 | 0 | 0 | 4 |
| Maxwell, 2017[63] | 1 | 1 | 0 | 1 | 1 | 0 | 1 | 5 |
| Mohammed, 2017[64] | 1 | 1 | 1 | 2 | 2 | 0 | 1 | 8 |
| Moore, 2016[65] | 1 | 1 | 1 | 1 | 2 | 0 | 1 | 7 |
| Moore, 2013[66] | 1 | 1 | 1 | 1 | 2 | 0 | 1 | 7 |
| Muhindo, 2019[67] | 1 | 1 | 0 | 1 | 2 | 0 | 1 | 6 |

| | | | | | | | | |
|---|---|---|---|---|---|---|---|---|
| Muhindo, 2020[68] | 1 | 1 | 1 | 1 | 2 | 0 | 1 | 7 |
| Murtaugh, 2020[69] | 1 | 1 | 1 | 2 | 2 | 0 | 1 | 8 |
| Ngo, 2013[70] | 1 | 1 | 0 | 1 | 2 | 1 | 1 | 6 |
| Pai, 2012[71] | 1 | 1 | 1 | 2 | 2 | 2 | 1 | 10 |
| Rocchetti, 2015[72] | 1 | 1 | 1 | 1 | 0 | 0 | 1 | 5 |
| Saunders, 2012[73] | 1 | 1 | 0 | 1 | 0 | 0 | 1 | 4 |
| Sullivan, 2021[74] | 0 | 1 | 1 | 2 | 1 | 0 | 1 | 6 |
| Tobin-West, 2013[75] | 1 | 1 | 1 | 1 | 1 | 0 | 1 | 6 |

| | | | | | | | | |
|---|---|---|---|---|---|---|---|---|
| Tucker, 2012[76] | 0 | 1 | 1 | 1 | 2 | 0 | 1 | 6 |
| Tucker, 2012[77] | 0 | 1 | 1 | 1 | 2 | 2 | 1 | 8 |
| Tucker, 2011[78] | 0 | 1 | 1 | 1 | 2 | 1 | 1 | 7 |
| Wang Cheng, 2019[79] | 0 | 1 | 1 | 0 | 2 | 2 | 1 | 7 |
| Williford, 2021[80] | 1 | 1 | 1 | 2 | 2 | 2 | 1 | 10 |
| Zhao, 2020[81] | 0 | 1 | 1 | 2 | 2 | 2 | 1 | 9 |

**Newcastle-Ottawa Scale – Case-control Studies**

| **Authors** | Is the case definition adequate?<br><br>a) yes, with independent validation<br><br>b) yes, eg record linkage or based on self reports<br><br>c) no description | Representativeness of the cases<br><br>a) consecutive or obviously representative series of cases<br><br>b) potential for selection biases or not stated | Selection of Controls<br><br>a) community controls<br><br>b) hospital controls<br><br>c) no description | Definition of Controls<br><br>a) no history of disease (endpoint)<br><br>b) no description of source | Comparability of cases and controls on the basis of the design or analysis<br><br>a) study controls for<br><br>_______________<br>(Select the most important factor.)<br><br>b) study controls for any additional factor Ø (This criteria could be modified to indicate specific control for a second important factor.) | Assessment of exposure<br><br>a) secure record (eg surgical records)<br><br>b) structured interview where blind to case/control status<br><br>c) interview not blinded to case/control status<br><br>d) written self report or medical record only<br>e) no description | Same method of ascertainment for cases and controls<br><br>a) yes<br>b) no. | Non-Response rate<br>a) same rate for both groups<br><br>b) non respondents described<br><br>c) rate different and no designation | Score |
|---|---|---|---|---|---|---|---|---|---|

| | | | | | | | | | |
|---|---|---|---|---|---|---|---|---|---|
| Joore, 2016[12] | 1 | 1 | 1 | 1 | 2 | 1 | 1 | 0 | 8 |

| Author | 1. Is there congruity between the stated philosophical perspective and the research methodology? | 2. Is there congruity between the research methodology and the research question or objectives? | 3. Is there congruity between the research methodology and the methods used to collect data? | 4. Is there congruity between the research methodology and the representation and analysis of data? | 5. Is there congruity between the research methodology and the interpretation of results? | 6. Is there a statement locating the researcher culturally or theoretically? | 7. Is the influence of the researcher on the research, and vice-versa, addressed? | 8. Are participants, and their voices, adequately represented? | 9. Is the research ethical according to current criteria or, for recent studies, and is there evidence of ethical approval by an appropriate body? | 10. Do the conclusions drawn in the research report flow from the analysis, or interpretation, of the data? |
|---|---|---|---|---|---|---|---|---|---|---|
| **Baker, 2015[82]** | Unclear | Y | Y | Y | Y | N | N | Y | Y | Y |
| **Balan, 2020[83]** | Unclear | Y | Y | Y | Y | N | N | Y | Y | Y |

| | | | | | | | | | | |
|---|---|---|---|---|---|---|---|---|---|---|
| **Bien, 2015[84]** | Unclear | Y | Y | Y | Y | N | N | Y | Unclear | Y |
| **Bradley, 2013[42]** | Unclear | Y | Y | Y | Y | N | N | Y | N | Y |
| **Jones, 2017[85]** | Y | Y | Y | Y | Y | N | N | Y | Y | Y |
| **Hottes, 2012[86]** | Unclear | Y | Y | Y | Y | N | N | Y | Y | Y |
| **Joore, 2017[87]** | Unclear | Y | Y | Y | Y | N | N | Y | N | Y |

| | | | | | | | | | | |
|---|---|---|---|---|---|---|---|---|---|---|
| **Knight, 2012[88]** | Y | Y | Y | Y | Y | Y | N | Y | Y | Y |
| **Lanier, 2014[16]** | Y | Y | Y | Y | Y | N | N | Y | N | Y |
| **McDonagh, 2019[89]** | Unclear | Y | Y | Y | Y | N | N | Unclear | N | Y |
| **Mullens, 2019[17]** | Unclear | Y | Y | Y | Y | N | N | Y | Y | Y |
| **Phrasisombath, 2012[90]** | Unclear | Y | Y | Y | Y | N | N | Y | Y | Y |
| **Scheim, 2016[91]** | Unclear | Y | Y | Y | Y | N | N | Y | Y | Y |
| **Slinkard, 2011[92]** | Unclear | Y | Y | Y | Y | N | N | Y | Y | Y |

| | | | | | | | | | | |
|---|---|---|---|---|---|---|---|---|---|---|
| **Sullivan, 2021[93]** | Unclear | Y | Y | Y | Y | N | N | Y | Y | Y |
| **Underhill, 2014[94]** | Unclear | Y | Y | Y | Y | N | N | Y | Y | Y |

94. Underhill K, Morrow KM, Colleran CM, Holcomb R, Operario D, Calabrese SK, et al. Access to healthcare, HIV/STI testing, and preferred pre-exposure prophylaxis providers among men who have sex with men and men who engage in street-based sex work in the US. PLoS ONE [Electronic Resource].9(11):e112425.
